# Localization of Abnormal Brain Regions in Parkinsonian Disorders: An ALE Meta-Analysis

**DOI:** 10.1101/2022.04.11.22273755

**Authors:** Elizabeth G. Ellis, Juho Joutsa, Jordan Morrison-Ham, Karen Caeyenberghs, Daniel T. Corp

## Abstract

Parkinsonism is a feature of several neurodegenerative disorders, including Parkinson’s disease (PD), progressive supranuclear palsy (PSP), corticobasal degeneration syndrome (CBS) and multiple system atrophy (MSA). Neuroimaging studies have yielded insights into parkinsonism; however it remains unclear whether there is a common neural substrate amongst disorders. The aim of the present meta-analysis was to identify consistent brain alterations in parkinsonian disorders (PD, PSP, CBS, MSA) both individually, and combined, to elucidate the shared substrate of parkinsonism. 33,505 studies were systematically screened following searches of MEDLINE Complete and Embase databases. A series of whole-brain activation likelihood estimation meta-analyses were performed on 126 neuroimaging studies (64 PD; 25 PSP; 18 CBS; 19 MSA) utilizing anatomical MRI, perfusion or metabolism positron emission tomography and single photon emission computed tomography. Abnormality of the caudate, thalamus, middle frontal and temporal gyri was common to all parkinsonian disorders. Localizations of commonly affected brain regions in individual disorders aligned with current diagnostic imaging markers, localizing the midbrain in PSP, putamen in MSA-parkinsonian variant and brainstem in MSA-cerebellar variant. Regions of the basal ganglia and precuneus were most commonly affected in PD, while CBS was characterized by caudate abnormality. To our knowledge, this is the largest meta-analysis of neuroimaging studies in parkinsonian disorders. Findings support the notion that parkinsonism may share a common neural substrate, independent of the underlying disease process, while also highlighting characteristic patterns of brain abnormality in each disorder.

## Introduction

Parkinsonism is a debilitating neurological syndrome characterized by bradykinesia, tremor, rigidity, and postural instability^1^. The syndrome is most often caused by Parkinson’s disease (PD) – the second most prevalent neurodegenerative disease worldwide, with incidence rates predicted to double in the next 20 years^2,3^. Parkinsonism can also be caused by several atypical parkinsonian disorders (APDs), including Progressive Supranuclear Palsy (PSP), Corticobasal Degeneration Syndrome (CBS), and Multiple System Atrophy (MSA)^2^. APDs are relatively rare and underdiagnosed, as the typical clinical characteristics can appear late in the course of the disease^4,5^. Identification of the neural substrates of these disorders is crucial to improving our understanding of their mechanisms^4,6,7^.

Given that parkinsonism is a shared symptom complex, identifying converging brain alterations across the disorders may clarify the common neural substrate of parkinsonism^8^. Traditionally, parkinsonism has been associated with dysfunction of the nigrostriatal tract^7,9^. Consequently, treatments to alleviate parkinsonism symptoms act on components of this circuit, including dopaminergic medications and deep brain stimulation^10,11^. Yet while these treatments have established efficacy in PD, APDs patients show little to no response^11,12^.

There are some characteristic imaging findings, including the “hot cross bun” sign of MSA, the “hummingbird” sign of PSP, and asymmetric cortical atrophy in CBS^13–15^, but sensitivity of these findings especially in the early phases of the diseases is relatively weak and shared neural substrates across disorder remain to be characterized. Inconsistencies in neuroimaging evidence, clinical heterogeneity, and methodological considerations (e.g., imaging modalities, study settings) may all contribute to heterogenous findings^6^.

Meta-analytic methods have demonstrated the ability to collate heterogeneous imaging findings to converge upon a small number of key brain regions in neurological disorders^16,17^. There is increasing evidence that symptoms share common neurobiological substrates^8,18^ but this hypothesis has not been tested across parkinsonian syndromes. Previous parkinsonian meta-analyses have focused on the individual parkinsonian disorders (for e.g.,^19–21^.

Therefore, the aim of the present study was to localize the neural substrates of parkinsonism, applying ALE meta-analysis to parkinsonian disorders, across imaging modalities. Meta-analyses were performed on both the individual parkinsonian disorders and in the combined cohort of ‘parkinsonism’ (PD + PSP + CBS + MSA).

## Methods

### Systematic Search

Embase and MEDLINE Complete databases were searched for studies of neuroimaging in patients with idiopathic PD, PSP, CBS, and MSA. Four systematic searches were conducted (i.e., one for each clinical diagnosis). No language or year limiters were applied. Studies were identified using a combination of key words relating to the diagnosis of focus, neuroimaging techniques, and imaging outcomes, for example: “parkinson* disease”, “magnetic resonance imaging”, “single photon emission computed tomography”, “atrophy” (for full syntax see Supplementary file 1). Studies were screened at title/abstract level using EndNote (Version X9) and Rayyan^22^ software, and full-text articles were screened in EndNote. Reference lists of all eligible studies were assessed for studies missed by the initial search.

#### Inclusion and Exclusion Criteria

To ensure objectivity in brain regions identified, studies must have: 1) reported whole-brain neuroimaging analysis^17^; 2) reported anatomical coordinates of significant brain abnormalities in stereotactic space; 3) provided clinical diagnosis information; 4) employed a healthy participant comparison group.

Exclusion criteria were: 1) studies using neurotransmitter or protein ligands and neuromelanin-sensitive MRI, excluded from the current analysis to avoid biased identification of regions dense in particular neurotransmitters, proteins, and melanin, respectively; 2) studies examining parkinsonism resulting from confirmed pathology (e.g., drug induced, familial, or a consequence of other disease); 3) studies not in English; 4) case studies, reviews and meta-analyses.

### Activation Likelihood Estimation Meta-Analyses

ALE meta-analyses were conducted as per the revised algorithm presented by Eickhoff et al. (2012), implementing a random-effects model^17^, refined permutation testing and corrections for multiple comparisons. Prior to ALE analyses, Talairach coordinates were transformed into MNI space using the Yale BioImage Suite converter^23^ (https://bioimagesuiteweb.github.io/webapp/mni2tal.html). Analyses were performed using GingerALE (v3.2; http://www.brainmap.org/ale/). In this revised algorithm each coordinate (‘foci’) is represented by a Gaussian probability distribution reflecting the underlying spatial uncertainty of each point. The width of each distribution is determined by the sample size of the study from which they were extracted^16^. Voxel-wise pooling of these probability distributions occurs to generate whole-brain ‘Modelled Activation’ (MA) maps - within which, each voxel represents the probability that a coordinate exists at that location (for e.g., the probability that volume loss occurred at that given voxel). The union of these individual MA maps was computed and referred to as the ‘ALE map’. To determine significance, the ALE map was then tested against the null distribution of 1000 simulated datasets with identical characteristics to the input (i.e., number of studies, sample sizes and number of foci) but with randomly distributed foci^17^. Within a cluster-level design, MA-values are distributed throughout the brain to determine which clusters of convergence from the included studies are significantly larger in their spatial extent than could be expected by chance.

Each analysis was performed with cluster-forming thresholds of *p* < .001 (uncorrected), 1000 permutations and a cluster-level inference threshold correcting for multiple comparisons with family-wise error rate (FWE) at *p* < .05^16^. Thresholds are in accordance with recommendations regarding both the sensitivity and specificity of cluster-level inference and corrections for multiple comparisons^24^. An exploratory threshold of uncorrected *p* < .001 with a minimum cluster extent threshold of 100mm^3^ was applied if contrasts were non-significant.

Prior to main analyses, we ran analyses separately within each neuroimaging modality (MRI, PET, SPECT), and performed ALE ‘contrast analyses’ to detect significant differences between the results of the modality-driven analyses (i.e., MRI vs PET, MRI vs SPECT, PET vs SPECT). Contrast analyses work by subtraction methods to provide a measure of the difference in convergence between two groups, revealing significant clusters unique to one of the two groups (for formulae see ^25^). Clusters were thresholded at uncorrected *p* < .001 to overcome potential limits of the false-discovery rate algorithm^24,26^, with 1000 permutations and a minimum cluster size of 100mm^3^. Results remained largely consistent between individual modality-driven analyses and main meta-analysis results (combined modalities). Only minor differences were identified (see Supplementary file 2) and therefore imaging modalities were combined for the following meta-analyses.

Analyses were performed in two contrasts. First, using coordinates of *decreased* volume, metabolism, or perfusion in patients compared to HC (i.e., parkinsonism < HC; PD < HC; PSP < HC; CBS < HC; MSA < HC). Second, using coordinates of *increased* volume, metabolism, or perfusion in each patient group compared to HC. In addition to the main MSA analyses, further analyses were conducted separating the MSA-Cerebellar (MSA-C) and MSA-Parkinsonian (MSA-P) clinical subtypes. Cluster regional labels for all analyses were defined using Harvard-Oxford cortical and subcortical structural atlases (FSL: https://fsl.fmrib.ox.ac.uk/fsl/fslwiki/Atlases).

## Results

### Systematic Literature Searches

A total of 33,505 articles were assessed for eligibility (Figure 1). After duplicate removal, 26,265 articles were screened at title/abstract level, and 1,416 at full-text. Ultimately, 126 studies met inclusion criteria (64 PD; 25 PSP; 18 CBS; 19 MSA; for all PRISMA flowcharts see Supplementary file 3). Searches were conducted in February 2019 (PD), July 2020 (PSP), May 2020 (CBS) and July 2020 (MSA). Meta-analyses for each disorder exceed the sample size requirements for the sufficient detection of power in ALE meta-analyses (i.e., each including more than 17 studies)^24,27^.

**Figure 1.**
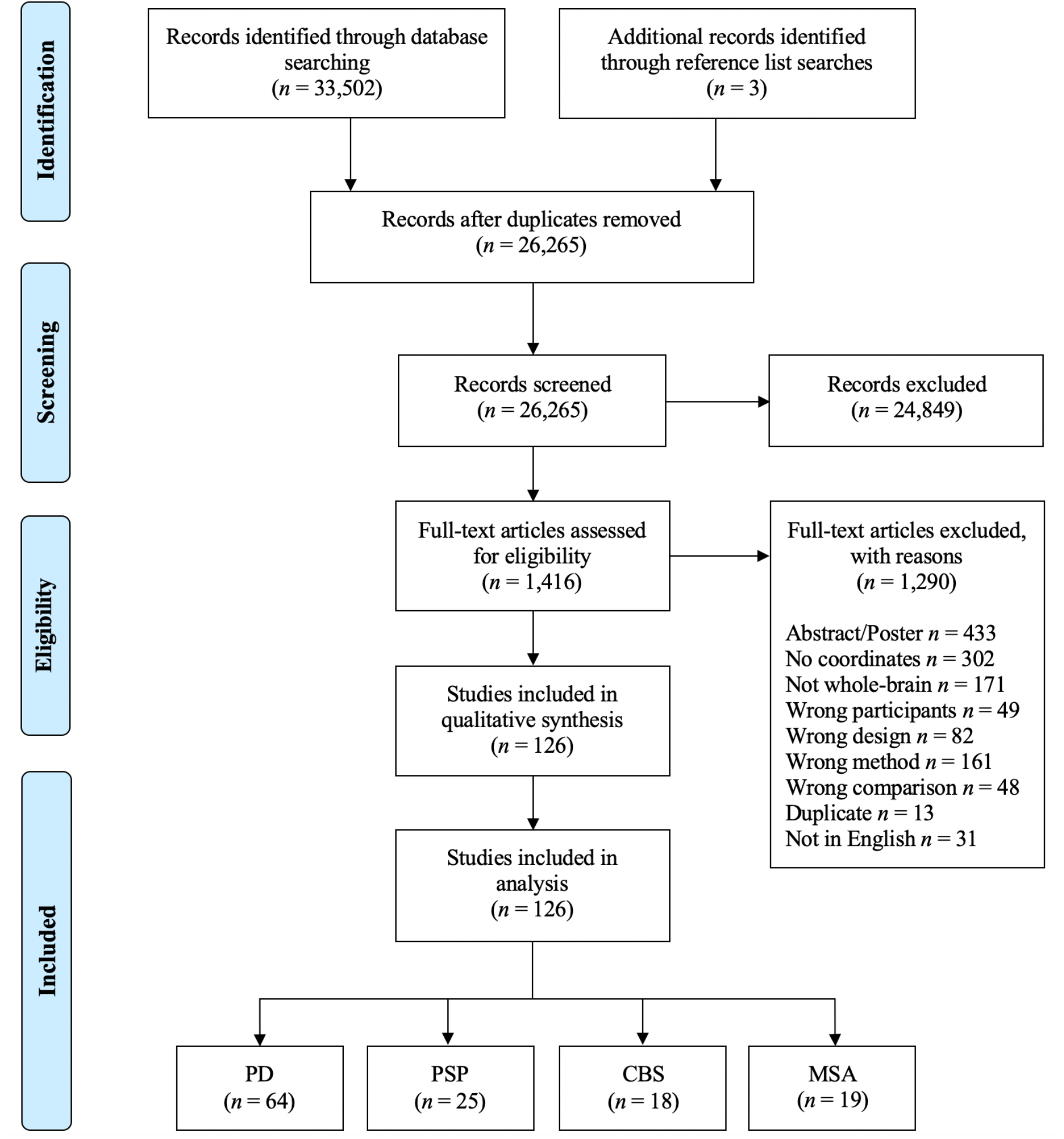
PRISMA Systematic Literature Search Decision Flowchart. PRISMA systematic literature search decision flowchart displaying results of the four searches combined. Adapted from: Moher et al., 2009.

### Activation Likelihood Estimation Meta-Analyses

Overall, the included studies comprised 7030 participants and 1961 coordinates of significant differences between patients and controls (Table 1). Imaging modalities of the included studies were whole-brain structural MRI, perfusion and metabolism positron emission tomography (PET) and single photon emission computed tomography (SPECT; for details of included studies see Supplementary file 4). Significant ALE meta-analysis results of all cohorts are detailed in Table 2. Figures are presented using uncorrected thresholds (*p* < .001) for visualization purposes^28,29^.

**Table 1.**
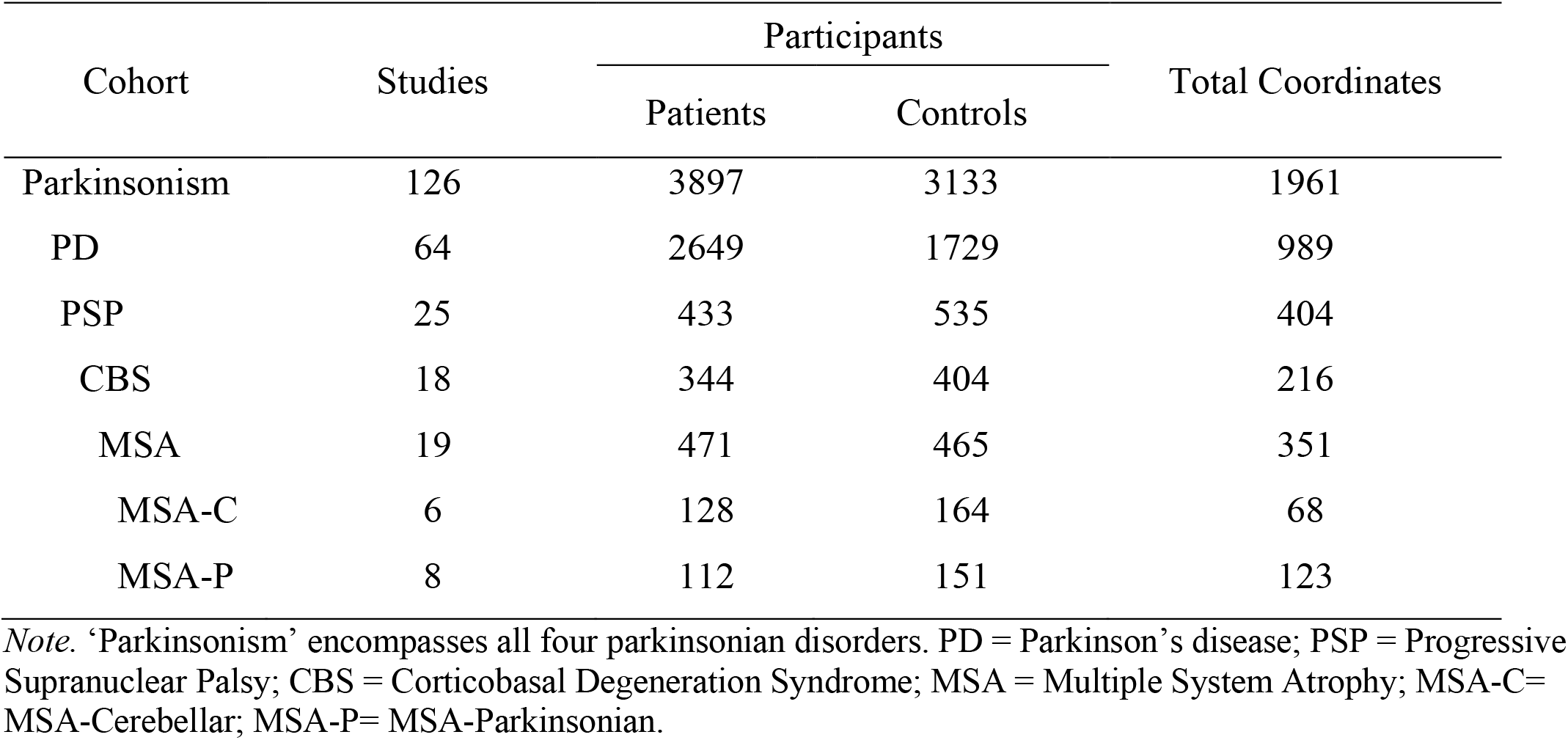
Cohorts Included in Meta-Analyses

| Cohort | Studies | Participants |  | Total Coordinates |
| --- | --- | --- | --- | --- |
|  |  | Patients | Controls |  |
| Parkinsonism | 126 | 3897 | 3133 | 1961 |
| PD | 64 | 2649 | 1729 | 989 |
| PSP | 25 | 433 | 535 | 404 |
| CBS | 18 | 344 | 404 | 216 |
| MSA | 19 | 471 | 465 | 351 |
| MSA-C | 6 | 128 | 164 | 68 |
| MSA-P | 8 | 112 | 151 | 123 |

*Note.* ‘Parkinsonism’ encompasses all four parkinsonian disorders. PD = Parkinson’s disease; PSP = Progressive Supranuclear Palsy; CBS = Corticobasal Degeneration Syndrome; MSA = Multiple System Atrophy; MSA-C= MSA-Cerebellar; MSA-P= MSA-Parkinsonian.

**Table 2.**
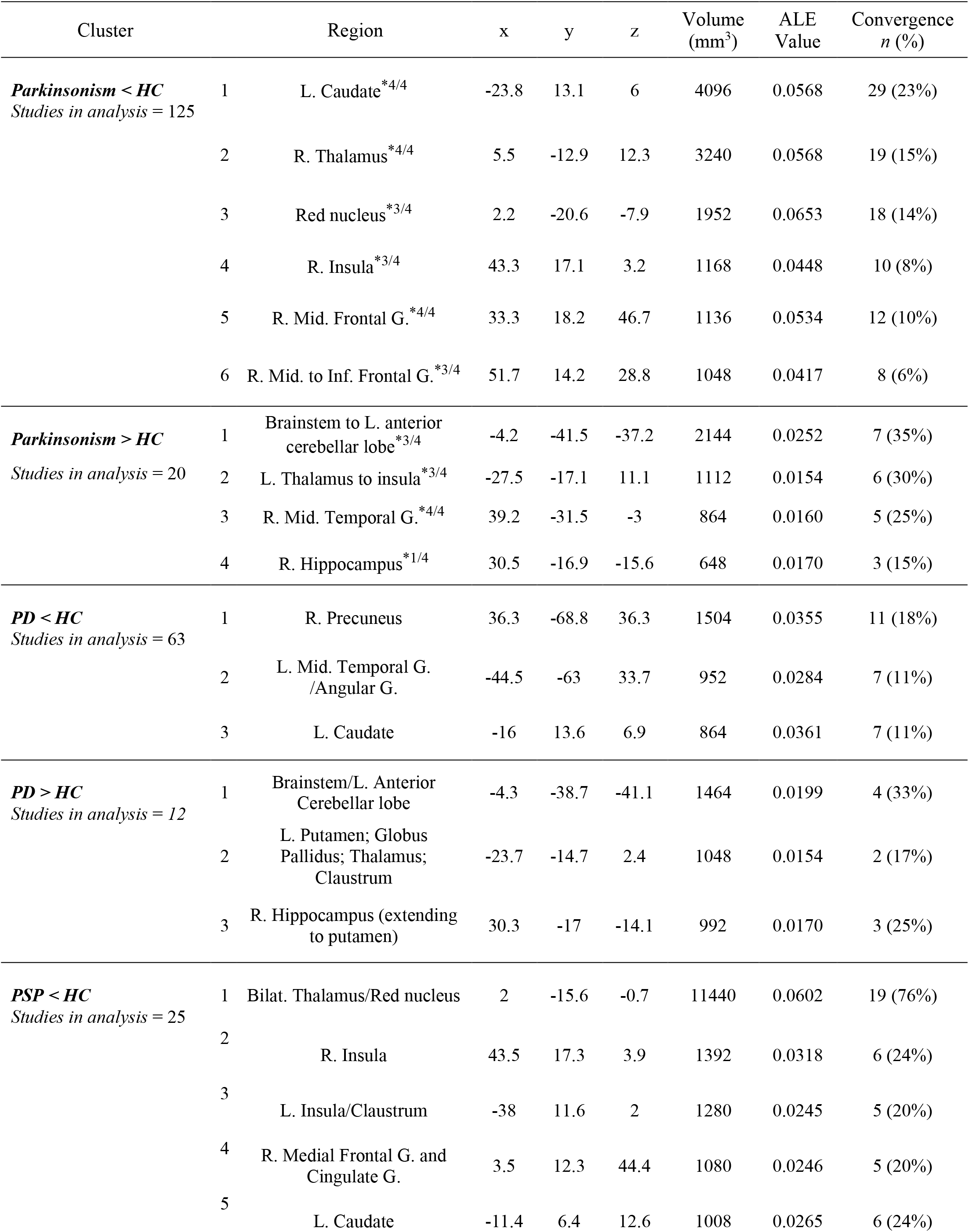
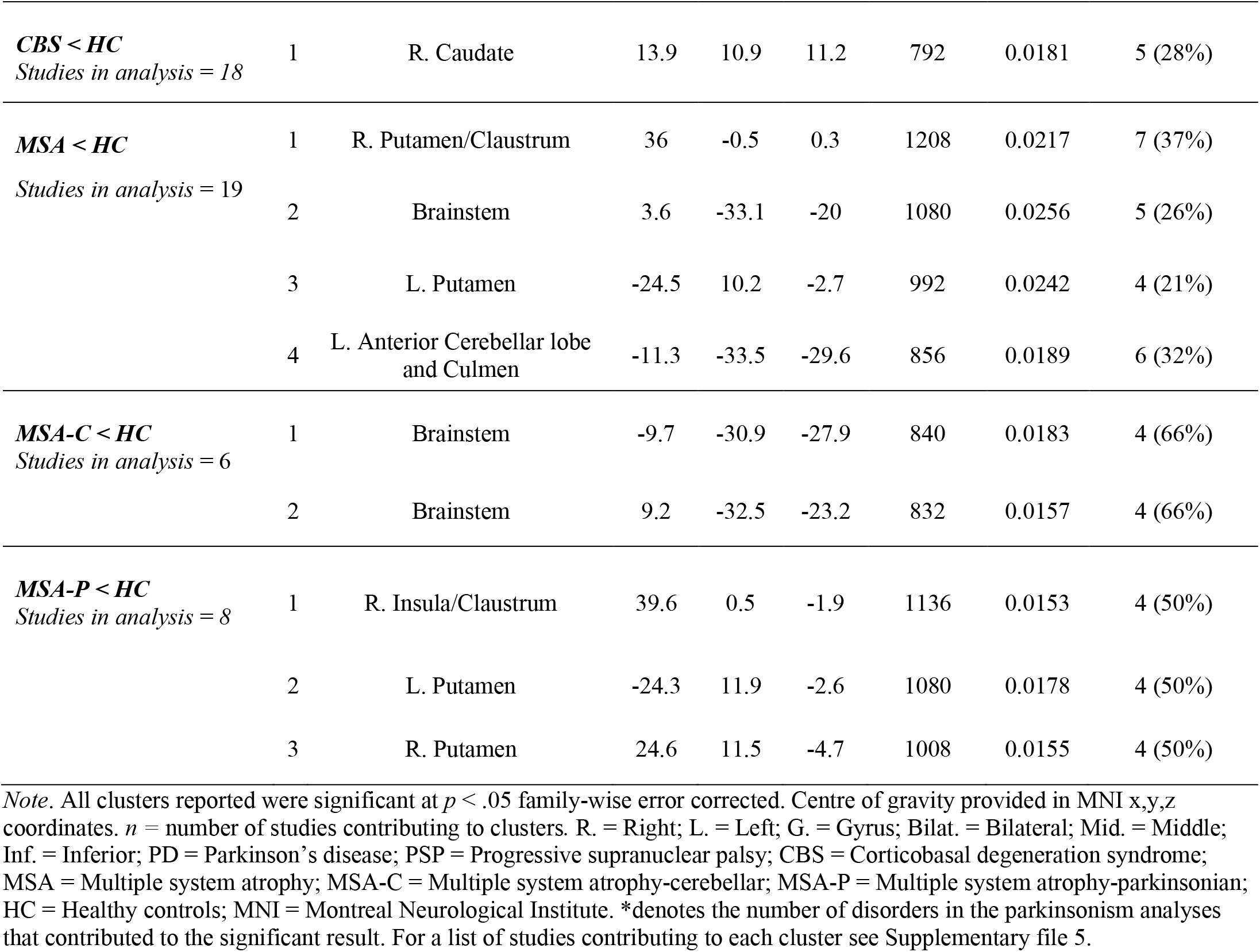
Consistent Regions of Abnormality in Parkinsonian Disorders

| Cluster |  | Region | x | y | z | Volume (mm<sup>3</sup>) | ALE Value | Convergence n (%) |
| --- | --- | --- | --- | --- | --- | --- | --- | --- |
| ***Parkinsonism < HC***<br>*Studies in analysis = 125* | 1 | L. Caudate <sup>*4/4</sup> | -23.8 | 13.1 | 6 | 4096 | 0.0568 | 29 (23%) |
|  | 2 | R. Thalamus <sup>*4/4</sup> | 5.5 | -12.9 | 12.3 | 3240 | 0.0568 | 19 (15%) |
|  | 3 | Red nucleus <sup>*3/4</sup> | 2.2 | -20.6 | -7.9 | 1952 | 0.0653 | 18 (14%) |
|  | 4 | R. Insula <sup>*3/4</sup> | 43.3 | 17.1 | 3.2 | 1168 | 0.0448 | 10 (8%) |
|  | 5 | R. Mid. Frontal G. <sup>*4/4</sup> | 33.3 | 18.2 | 46.7 | 1136 | 0.0534 | 12 (10%) |
|  | 6 | R. Mid. to Inf. Frontal G. <sup>*3/4</sup> | 51.7 | 14.2 | 28.8 | 1048 | 0.0417 | 8 (6%) |
| ***Parkinsonism > HC***<br>*Studies in analysis = 20* | 1 | Brainstem to L. anterior cerebellar lobe <sup>*3/4</sup> | -4.2 | -41.5 | -37.2 | 2144 | 0.0252 | 7 (35%) |
|  | 2 | L. Thalamus to insula <sup>*3/4</sup> | -27.5 | -17.1 | 11.1 | 1112 | 0.0154 | 6 (30%) |
|  | 3 | R. Mid. Temporal G. <sup>*4/4</sup> | 39.2 | -31.5 | -3 | 864 | 0.0160 | 5 (25%) |
|  | 4 | R. Hippocampus <sup>*1/4</sup> | 30.5 | -16.9 | -15.6 | 648 | 0.0170 | 3 (15%) |
| ***PD < HC***<br>*Studies in analysis = 63* | 1 | R. Precuneus | 36.3 | -68.8 | 36.3 | 1504 | 0.0355 | 11 (18%) |
|  | 2 | L. Mid. Temporal G. /Angular G. | -44.5 | -63 | 33.7 | 952 | 0.0284 | 7 (11%) |
|  | 3 | L. Caudate | -16 | 13.6 | 6.9 | 864 | 0.0361 | 7 (11%) |
| ***PD > HC***<br>*Studies in analysis = 12* | 1 | Brainstem/L. Anterior Cerebellar lobe | -4.3 | -38.7 | -41.1 | 1464 | 0.0199 | 4 (33%) |
|  | 2 | L. Putamen; Globus Pallidus; Thalamus; Claustrum | -23.7 | -14.7 | 2.4 | 1048 | 0.0154 | 2 (17%) |
|  | 3 | R. Hippocampus (extending to putamen) | 30.3 | -17 | -14.1 | 992 | 0.0170 | 3 (25%) |
| ***PSP < HC***<br>*Studies in analysis = 25* | 1 | Bilat. Thalamus/Red nucleus | 2 | -15.6 | -0.7 | 11440 | 0.0602 | 19 (76%) |
|  | 2 | R. Insula | 43.5 | 17.3 | 3.9 | 1392 | 0.0318 | 6 (24%) |
|  | 3 | L. Insula/Clastrum | -38 | 11.6 | 2 | 1280 | 0.0245 | 5 (20%) |
|  | 4 | R. Medial Frontal G. and Cingulate G. | 3.5 | 12.3 | 44.4 | 1080 | 0.0246 | 5 (20%) |
|  | 5 | L. Caudate | -11.4 | 6.4 | 12.6 | 1008 | 0.0265 | 6 (24%) |
| **CBS < HC**<br>*Studies in analysis = 18* | 1 | R. Caudate | 13.9 | 10.9 | 11.2 | 792 | 0.0181 | 5 (28%) |
| **MSA < HC**<br>*Studies in analysis = 19* | 1 | R. Putamen/Clastrum | 36 | -0.5 | 0.3 | 1208 | 0.0217 | 7 (37%) |
|  | 2 | Brainstem | 3.6 | -33.1 | -20 | 1080 | 0.0256 | 5 (26%) |
|  | 3 | L. Putamen | -24.5 | 10.2 | -2.7 | 992 | 0.0242 | 4 (21%) |
|  | 4 | L. Anterior Cerebellar lobe and Culmen | -11.3 | -33.5 | -29.6 | 856 | 0.0189 | 6 (32%) |
| **MSA-C < HC**<br>*Studies in analysis = 6* | 1 | Brainstem | -9.7 | -30.9 | -27.9 | 840 | 0.0183 | 4 (66%) |
|  | 2 | Brainstem | 9.2 | -32.5 | -23.2 | 832 | 0.0157 | 4 (66%) |
| **MSA-P < HC**<br>*Studies in analysis = 8* | 1 | R. Insula/Clastrum | 39.6 | 0.5 | -1.9 | 1136 | 0.0153 | 4 (50%) |
|  | 2 | L. Putamen | -24.3 | 11.9 | -2.6 | 1080 | 0.0178 | 4 (50%) |
|  | 3 | R. Putamen | 24.6 | 11.5 | -4.7 | 1008 | 0.0155 | 4 (50%) |

*Note.* All clusters reported were significant at $p < .05$ family-wise error corrected. Centre of gravity provided in MNI x,y,z coordinates. $n$ = number of studies contributing to clusters. R. = Right; L. = Left; G. = Gyrus; Bilat. = Bilateral; Mid. = Middle; Inf. = Inferior; PD = Parkinson's disease; PSP = Progressive supranuclear palsy; CBS = Corticobasal degeneration syndrome; MSA = Multiple system atrophy; MSA-C = Multiple system atrophy-cerebellar; MSA-P = Multiple system atrophy-parkinsonian; HC = Healthy controls; MNI = Montreal Neurological Institute. \*denotes the number of disorders in the parkinsonism analyses that contributed to the significant result. For a list of studies contributing to each cluster see Supplementary file 5.

#### Meta-Analysis of Parkinsonism

Six significant clusters were identified in the parkinsonism < HC analysis (*p* < .05 FWE-corrected) (Figure 2, Table 2). The left caudate, right thalamus and right middle frontal gyrus clusters were driven by coordinates of abnormality from all four parkinsonian disorders (Table 2). The remaining three clusters were driven by coordinates from three of the four parkinsonian disorders.

**Figure 2.**
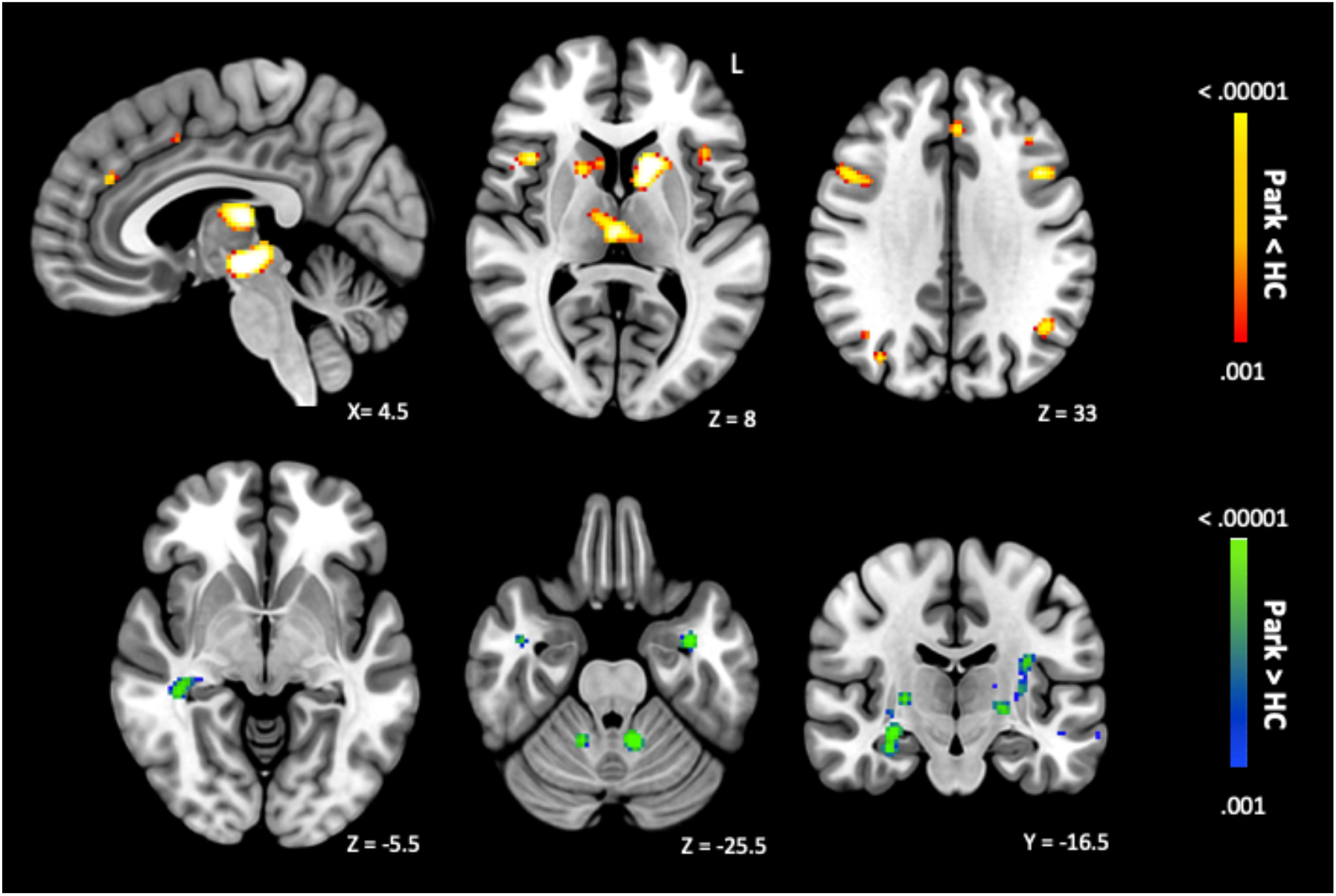
Meta-Analysis Findings in Parkinsonism. Abnormality of the caudate, thalamus, middle frontal and middle temporal gyri was identified across all four parkinsonian disorders. Note that figures are presented using uncorrected *p* thresholds (*p* < .001) for visualization purposes. *Red to white* = parkinsonism < HC (top row). *Blue to green* = parkinsonism > HC (bottom row). Park = parkinsonism; HC = healthy controls.

Four significant clusters were identified in the parkinsonism > HC contrast (Figure 2, bottom row; Table 2). The right middle temporal gyrus cluster was driven by coordinates of abnormality from all four parkinsonian disorders (Table 2).

#### Parkinson’s Disease Meta-Analysis

Three significant clusters were identified by the PD < HC meta-analysis. Clusters were centered upon the right precuneus, left middle temporal gyrus and caudate (*p* < .05 FWE-corrected) (Figure 3A, first row; Table 2).

**Figure 3.**
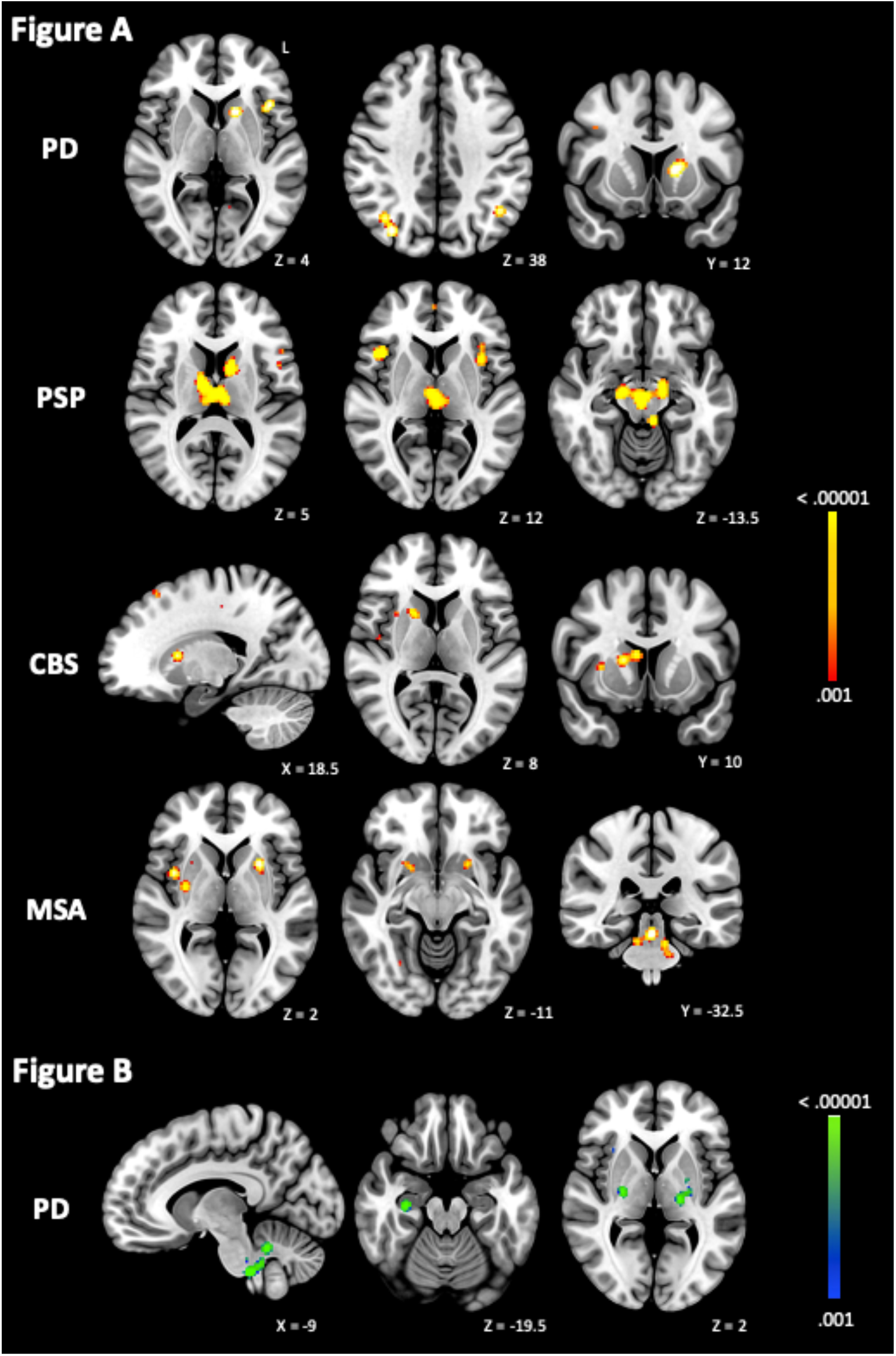
Meta-Analysis Results in Individual Parkinsonian Disorders. Figure 3.A: patients < HC. The precuneus was the most commonly affected brain region in PD, whilst alteration of the red nucleus was characteristic of PSP patients. The caudate and the putamen were the most commonly implicated regions in patients with CBS and MSA, respectively. Figure 3.B: PD > HC. Structures of the brainstem, basal ganglia and hippocampus were consistent regions of increases in PD patients compared to controls. Note that figures are presented using uncorrected *p* thresholds (*p* < .001) for visualization purposes. PD = Parkinson’s disease; PSP = Progressive supranuclear palsy; CBS = Corticobasal degeneration syndrome; MSA = Multiple system atrophy.

In the opposite contrast, PD > HC, three clusters were significant within the brainstem/anterior cerebellar lobe, basal ganglia and right hippocampus (Figure 3B; Table 2).

#### Progressive Supranuclear Palsy Meta-Analysis

Five significant clusters were identified by the PSP < HC analysis (*p* < .05 FWE-corrected). Clusters were centered upon the bilateral thalamus/red nucleus, right and left insula, right medial frontal gyrus and left caudate (Figure 3A, second row; Table 2). No clusters survived thresholding in the opposite contrast (PSP > HC). Using an exploratory threshold (*p* < .001, uncorrected) two clusters were identified (see Supplementary file 6).

#### Corticobasal Degeneration Syndrome Meta-Analysis

One significant cluster was identified in the CBS < HC analysis, upon the right caudate (*p* < .05 FWE-corrected) (Figure 3, third row; Table 2). As only one study reported coordinates for the CBS > HC contrast^30^ no analysis was performed.

#### Multiple System Atrophy Meta-Analysis

The final ALE meta-analyses were performed within MSA. The MSA < HC meta-analysis identified four significant clusters (*p* < .05 FWE-corrected), with the largest cluster centered upon the right putamen/claustrum (Figure 3, bottom row; Table 2). No clusters survived thresholding in the MSA > HC contrast. Using an exploratory threshold (*p* < .001, uncorrected) ten clusters were identified (see Supplementary file 6). Results across all MSA patients implicated regions of the basal ganglia, brainstem, and cerebellum. However, when separating the MSA-C and MSA-P subtypes, distinct brain regions previously associated with their clinical presentations were localized.

***MSA-C***. Two significant clusters were identified in the MSA-C < HC analysis (*p* < FWE-corrected), both within the brainstem (Figure 4, top row). As only one study reported coordinates for the MSA-C > HC contrast, no analysis was performed. ***MSA-P***. Three significant clusters were identified in the MSA-P < HC analysis, upon the right insula, left and right putamen (*p* < .05 FWE-corrected; Figure 4, bottom row). No studies reported coordinates for the MSA-P > HC contrast.

**Figure 4:**
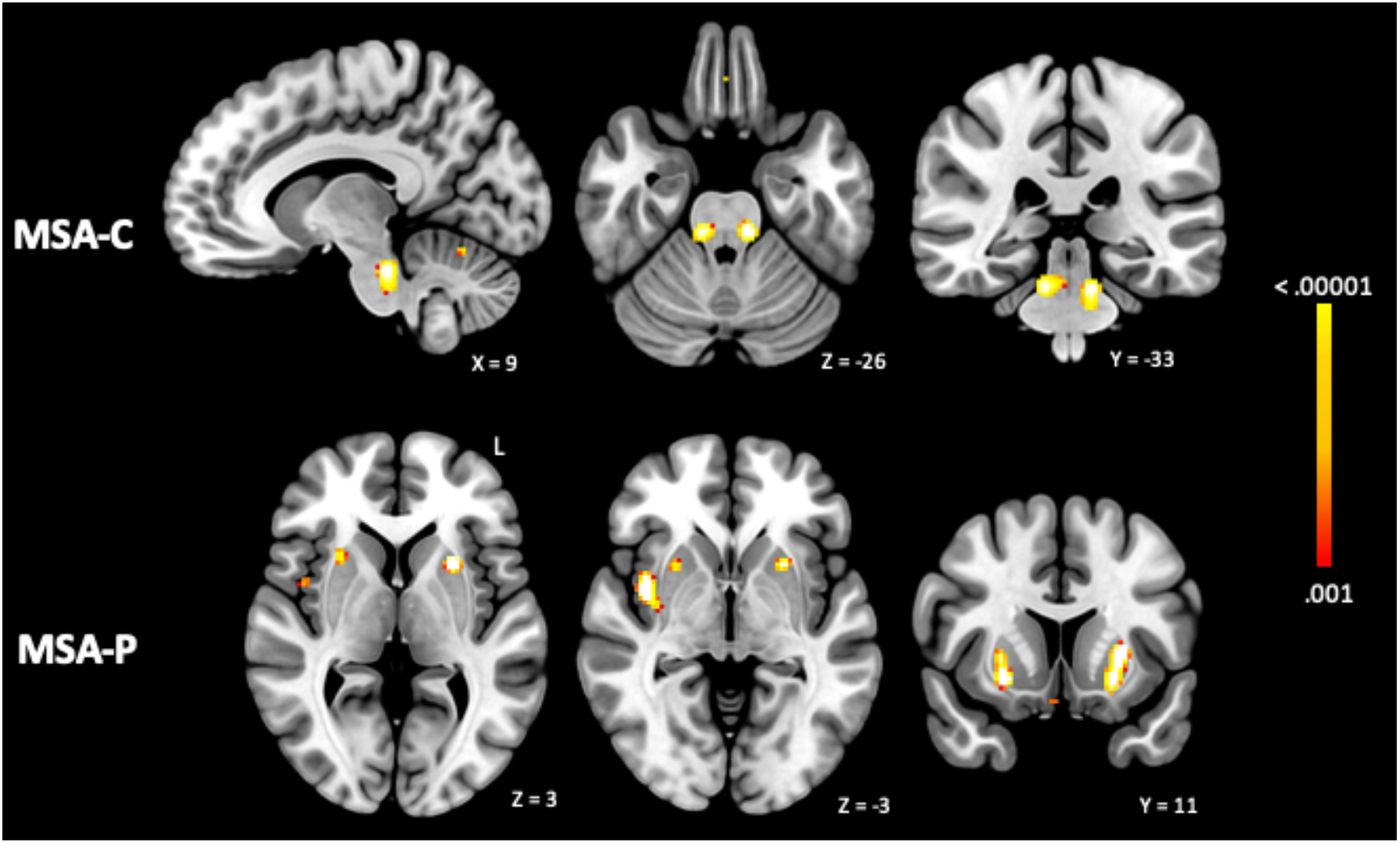
Multiple system atrophy subtype meta-analysis results. When divided by subtype, distinct regions of consistent brain alteration were identified for MSA-C and MSA-P patients, in alignment with their clinical presentation. Note that figures are presented using uncorrected *p* thresholds (*p* < .001) for visualization purposes. MSA-C = Multiple system atrophy-Cerebellar; MSA-P = Multiple system atrophy-Parkinsonian

## Discussion

The present study conducted a series of whole-brain ALE meta-analyses to identify consistent regions of brain abnormality across parkinsonian disorders. Reconciling previously inconsistent neuroimaging findings, abnormality of the caudate, thalamus, middle frontal and temporal gyri was common to all parkinsonian disorders. Patterns of consistent brain alteration identified in PSP and MSA aligned with clinical imaging markers, whilst the precuneus and caudate were the most commonly implicated regions in PD and CBS, respectively. Findings support the notion of a shared neural substrate for overlapping symptoms amongst these disorders, whilst also highlighting the disorders’ individual patterns of brain alteration.

### Consistent Regions of Abnormality Across Parkinsonian Disorders

Abnormality of the caudate, thalamus, right middle frontal and temporal gyri was common across all parkinsonian disorders. Identification of this set of shared brain abnormalities may support the notion that symptoms share a common neural substrate. These regions may be involved with both the overlapping motor and non-motor symptoms amongst disorders. For example, the thalamus, a hub of connections between cortical and subcortical structures, has an established role within basal ganglia circuitry, associated with a range of functions including motor control^31^. However, the caudate has been more commonly associated with cognitive and associative functions, alongside its sensorimotor connections^32^. Both the caudate and thalamus have been localized in previous meta-analyses as regions commonly affected within individual parkinsonian disorders (for e.g.,^6,19,20^).

Similarly, although the middle frontal and temporal gyri do not have an established role in the parkinsonism syndrome, both regions have been implicated in studies of individual disorders (for e.g.,^33,34^). These cortical regions have also been linked to the reorientation of attention^35^ and freezing of gait^36^, respectively. The role that the middle frontal gyrus plays in gait disturbances in parkinsonian patients warrants further examination, as the previous association was identified with *hypo*metabolism of the region, contradictory to our finding of consistent increases within the region.

The fact that these regions were found to be consistently abnormal across parkinsonian disorders may suggest that they form part of a shared substrate for shared symptoms. However, we cannot rule out that these sites of abnormality may be shared across disorders because of particular vulnerability to degeneration as opposed to as a result of their direct involvement in symptom generation^19,37^. Though this must be acknowledged, our hypothesis that these common regions may be involved in the shared substrate may be supported by the previous localizations of these regions in meta-analyses of the individual disorders (for e.g.,^6,20,21,26^). Furthermore, it could be that these regions are not discretely involved in symptom generation but part of a broader network of regions^18,38^.

### Consistent Regions of Abnormality Within Parkinsonian Disorders Parkinson’s Disease

Our PD meta-analyses further confirmed the neuroanatomical heterogeneity of PD, with a maximum of 11 studies found to report alteration to the same brain region. This evidence, in alignment with previous literature, suggests a number of different brain regions are involved in PD^7,39,40^. PD was characterized by alteration to the precuneus, temporal/angular gyri, and left caudate, consistent with previous meta-analyses^6,20,21^. Although the role of the precuneus in PD is not yet fully established, the region has been associated with cognitive deficits in PD patients^41^ as well as agency of movement and motor integration^42,43^, domains known to be disrupted in PD^44^.

Identification of consistent abnormality of the left middle temporal gyrus is congruent with its association with freezing of gait in patients with PD^36,45^. Of note, hypometabolism of this region has been shown to moderate the efficacy of deep brain stimulation in treating freezing of gait^36^. Further exploration within the context of parkinsonism motor symptoms may be warranted to examine the region’s potential to moderate the therapeutic effect across other motor symptoms. Finally, localization of the left caudate is in agreement with Albrecht et al.^6^, reporting consistent hypometabolism of particularly the left caudate to be associated with motor impairment in PD patients.

### Progressive Supranuclear Palsy

PSP was characterized by midbrain abnormality, consistent with previous meta-analyses^20,21^. This alteration is hallmark PSP pathology^2,46,47^, and proposed to be an imaging marker for differential diagnosis^26,48^. In addition, the consistent alteration to the basal ganglia and thalamus may align with the postural instability and ocular motor dysfunction often exhibited by PSP patients^49,50^.

### Corticobasal Degeneration Syndrome

The right caudate was the most commonly implicated region in CBS, as observed in previous meta-analyses^19,21^ and histopathological examinations^51^. Localization of only the right and not left caudate may in part be explained by the distinctive asymmetry of CBS symptomology and atrophy (for e.g.,^30,52–54^). Previous meta-analyses have identified more distributed alterations in CBS, involving the thalamus, insula and multiple cortical sites^19,21^ yet our meta-analysis identified a single significant region. There are two possible explanations for this. First, previous meta-analyses applied less stringent statistical thresholding; and second, both prior meta-analyses included only voxel-based morphometry studies (a structural MRI based technique used to identify volumetric change; ^55^). Our analysis including only structural-MRI studies in CBS was non-significant and required the use of exploratory uncorrected thresholds to obtain significant findings. Thus, it is likely the use of strict thresholding that is the determining factor in the number of brain regions found to be significant.

### Multiple System Atrophy

MSA was characterized by consistent alteration within the basal ganglia, brainstem and cerebellum, in agreement with previous meta-analyses^21,56,57^. Localizations align with the clinical subtypes, MSA-P and MSA-C, linked to the basal ganglia and cerebellum, respectively^58^. In addition, identification of robust abnormality within the basal ganglia and brainstem is in alignment with imaging markers used for differential diagnosis, including the distinctive MSA ‘hot cross bun’ sign within the pons^13^. Beyond imaging markers, disruption to cerebellar and brainstem structures is consistent with the cerebellar and autonomic dysfunction used to clinically distinguish MSA patients from other parkinsonian disorders^13^. Importantly, this was the first ALE meta-analysis to specifically examine the MSA-C subtype, allowing for examination of the distinct alteration patterns of MSA-C and MSA-P. Results are congruent with each subtype’s prominent presentation, with consistent abnormality of the brainstem of MSA-C and the putamen of MSA-P patients^15,58^.

### Localization of Brain Networks Versus Brain Regions

Whilst several of our meta-analyses demonstrated high convergence, there were still a number of studies that did not contribute to our ALE findings. For example, a maximum of 11 of 64 studies converged in PD. Although these findings may suggest a lack of reproducibility, it is also possible that the remainder of these studies’ coordinates are still functionally connected to the clusters that we identified. Complex symptoms which previously were ineffectively localized to discrete brain regions have been shown to more effectively localize to distributed brain networks^18,59,60^. Working with the principal of diaschisis, by which we know damaged brain regions may influence the functioning of other distal but connected regions which are responsible for symptom generation, a technique termed ‘lesion network mapping’ leverages a large database of normative functional MRI data to identify brain regions functionally connected to previously reported sites of abnormality^61^. Relevant to the present paper, Joutsa et al.^62^ demonstrated that brain lesions causing parkinsonism (secondary parkinsonism) are distributed across the brain, yet are commonly connected to the claustrum, basal ganglia, midbrain and cerebellum. Regions which were also implicated by our ALE meta-analyses, reinforcing that they may form part of the neural substrate of parkinsonism. Although our findings were derived from coordinates in idiopathic parkinsonian disorders (without causal brain lesions), it is also possible to map the connectivity of such peak coordinates^18^. It is plausible that the use of such a network-based paradigm may extend upon the convergence achieved by the present meta-analysis.

### Limitations

Our meta-analysis has a number of limitations that should be discussed. First, most of the included studies did not have diagnoses confirmed by autopsy. Due to the poor sensitivity of clinical APDs diagnoses and overdiagnosis of PD^4^, misdiagnoses within the published studies is possible. Second, ALE results are driven by coordinates of statistically significant differences between patients and controls. Whether patient differences reach significance in each study will have been influenced by individual study characteristics (e.g., image processing, statistical thresholds applied), choices which are not uniform across imaging literature. Whilst the ALE technique is able to partially control for variance in study methodologies, it is not able to do so completely. Third, without the use of neurodegenerative control conditions, it is possible that some of the regions identified by our parkinsonism analyses may be commonly abnormal across disorders because of vulnerability of certain regions to degeneration rather than due to their specific involvement in the production of shared symptoms^1,63^.

## Conclusion

To our knowledge, this is the largest whole-brain meta-analysis of parkinsonian disorders. Our meta-analyses reconciled previously inconsistent neuroimaging evidence, demonstrating that the caudate, thalamus, middle frontal and temporal gyri are commonly affected across parkinsonian disorders. Localizations aligned with current diagnostic imaging markers, isolating the midbrain in PSP, the brainstem in MSA, and the putamen and brainstem in MSA-P and -C, respectively. PD was defined by precuneus and basal ganglia alterations, whilst CBS was characterized by caudate abnormality. Findings support the notion that shared symptoms across these disorders may have a common neural substrate, independent of the underlying disease process, while also highlighting the individual characteristic patterns of brain abnormality in each disorder.

## Data Availability

The data that support the findings of this study are available from the corresponding author upon request. Code used via Ginger ALE version 3.2 is available through http://brainmap.org.

## Acronyms

PD: Parkinson’s disease
PSP: Progressive supranuclear palsy
CBS: Corticobasal degeneration syndrome
MSA: Multiple System Atrophy
APDs: Atypical parkinsonian disorders
ALE: Activation Likelihood Estimation
MRI: magnetic resonance imaging
PET: positron emission tomography
SPECT: single photon emission computed tomography

## Author Contributions

Elizabeth G. Ellis: 1abc, 2abc, 2ab

Juho Joutsa: 1b 2c 3b

Jordan Morrison-Ham: 2c 3b

Karen Caeyenberghs: 2c 3b

Daniel T. Corp: 1abc, 2ac, 3ab

1. **Research project**. A. Conception; B. Organization; C. Execution.
2. **Statistical analysis**. A. Design; B. Execution; C. Review/Critique.
3. **Manuscript**. A. Writing of the first draft; B. Review/Critique.

