## Supplementary file 1 for "Localization of Abnormal Brain Regions in Parkinsonian Disorders: An ALE Meta-Analysis"

**Supplementary file 1. Search Syntax**

**Table S1.1** *Systematic Search Syntax for Individual Parkinsonian Disorders*

| Database | Syntax |
| --- | --- |
| Embase | #1 AND #2 AND #3 |
| #1 | ('mri':ti OR 'magnetic resonance imaging':ti OR 'vbm':ti OR 'voxel based morpho*':ti OR 'spect':ti OR 'single photon emission computed tomography':ti OR 'positron emission tomography':ti OR 'pet':ti OR 'hypometabolism':ti OR 'hypoperfusion':ti OR 'mri':ab OR 'magnetic resonance imaging':ab OR 'vbm':ab OR 'voxel based morpho*':ab OR 'spect':ab OR 'single photon emission computed tomography':ab OR 'positron emission tomography':ab OR 'pet':ab OR 'hypometabolism':ab OR 'hypoperfusion':ab OR 'nuclear magnetic resonance imaging'/de OR 'positron emission tomography'/de OR 'single photon emission computed tomography'/de OR ((atrophy NEAR/9 (brain OR matter OR cort* OR neuro*)):ti,ab)) |
| #2a | ('parkinson* disease':ti OR 'parkinson* disease':ab OR 'parkinson disease'/de) |
| #2b | (‘Corticobasal degeneration':ti OR 'CBD':ti OR 'cortical basal ganglionic degeneration':ti OR 'corticobasal syndrome':ti OR 'CBS':ti OR 'corticodentatonigral degeneration with neuronal achromasia':ti OR 'corticonigral degeneration':ti OR 'Corticobasal degeneration':ab OR 'CBD':ab OR 'cortical basal ganglionic degeneration':ab OR 'corticobasal syndrome':ab OR 'CBS':ab OR 'corticodentatonigral degeneration with neuronal achromasia':ab OR 'corticonigral degeneration':ab OR 'corticobasal degeneration'/de) |
| #2c | ('multiple system* atrophy':ti OR 'msa':ti OR 'msa-p':ti OR 'msa-c':ti OR 'olivoponto-cerebellar atrophy':ti OR 'opca':ti OR 'striatonigral degeneration':ti OR 'snd':ti OR 'shy–drager syndrome':ti OR 'multisystem-atrophy':ti OR 'multi-system atrophy':ti OR 'multiple system* atrophy':ab OR 'msa':ab OR 'msa-p':ab OR 'msa-c':ab OR 'olivoponto-cerebellar atrophy':ab OR 'opca':ab OR 'striatonigral degeneration':ab OR 'snd':ab OR 'shy–drager syndrome':ab OR 'multisystem-atrophy':ab OR 'multi-system atrophy':ab OR 'multiple system atrophy'/de) |
| #2d | ('progressive supranuclear pals*':ti,ab OR 'psp':ti,ab OR 'steele-richardson-olszewski':ti,ab OR 'steele richardson olszewski':ti,ab OR 'richardson syndrome':ti,ab OR 'progressive supranuclear palsy'/de) |
| #3 | [embase]/lim |
| Medline Complete | #1 AND #2 |
| #1 | ( ( TI "MRI" OR TI "magnetic resonance imaging" OR AB "MRI" OR AB "magnetic resonance imaging" OR MH “magnetic resonance imaging” OR TI “VBM” OR AB “VBM” OR TI “voxel based morpho*” OR AB “voxel based morpho*” OR TI “SPECT” OR AB “SPECT” OR TI “single photon emission computed tomography” OR AB “single photon emission computed tomography” OR MH “Tomography, Emission-Computed, Single-Photon” OR TI “positron emission tomography” OR AB “positron emission tomography” OR TI “PET” OR AB “PET” OR MH “Positron-Emission Tomography” OR TI “hypometabolism” OR AB “hypometabolism” OR TI “hypoperfusion” OR AB “hypoperfusion” ) OR ( ( TI (atrophy) N9 (brain OR matter OR cort* or neuro*) ) OR ( AB (atrophy) N9 (brain OR matter OR cort* or neuro*) ) ) |
| #2a | (TI "parkinson* disease" OR AB "parkinson* disease" OR MH "parkinson disease" ) |
| #2b | (TI "Corticobasal degeneration" OR TI "CBD"OR TI "cortical basal ganglionic degeneration" OR TI "corticobasal syndrome" OR TI "CBS" OR TI "corticodentatonigral degeneration with neuronal achromasia" OR TI "corticonigral degeneration" OR AB "Corticobasal degeneration" OR AB "CBD" OR AB "cortical basal ganglionic degeneration" OR AB "corticobasal syndrome" OR AB "CBS" OR AB "corticodentatonigral degeneration with neuronal achromasia" OR AB "corticonigral degeneration" OR MH "corticobasal degeneration") |
| #2c | (TI "multiple system* atrophy" OR AB "multiple system* atrophy" OR TI "MSA" OR AB "MSA" OR TI "Olivoponto-cerebellar atrophy" OR AB "Olivoponto-cerebellar atrophy" OR TI "OPCA" OR AB "OPCA" OR TI "striatonigral degeneration" OR AB "striatonigral degeneration" OR TI "SND" OR AB "SND" OR TI "MSA-P" OR AB "MSA-P" OR TI "MSA-C" OR AB "MSA-C" OR TI "multisystem-atrophy" OR AB "multisystem-atrophy" OR TI "multi-system atrophy" OR AB "multi-system atrophy" OR TI "Shy–Drager syndrome" OR AB "Shy–Drager syndrome" OR MH "multiple system atrophy") |
| #2d | (TI "Progressive supranuclear pals*" OR TI "Steele-Richardson-Olszewski" OR TI "Steele Richardson Olszewski" OR TI "Richardson syndrome" OR TI "PSP" OR AB "Progressive supranuclear pals*" OR AB "Steele-Richardson-Olszewski" OR AB "Steele Richardson Olszewski" OR AB "Richardson syndrome" OR AB "PSP" OR MH "progressive supranuclear palsy") |
