## Supplementary file 2 for "Localization of Abnormal Brain Regions in Parkinsonian Disorders: An ALE Meta-Analysis"

**Supplementary file 2. Imaging Modality Driven Meta-Analysis Results**

**Imaging Modality Meta-analyses**

Results of the meta-analyses subdivided by imaging modality were largely consistent with the main analysis results (combined modalities). To detect any significant differences between imaging modality meta-analyses, ALE contrast analyses were conducted. Only one contrast was significant; clusters within the middle temporal gyrus, inferior frontal gyrus and left caudate were found to be specific to PET imaging compared to MRI (Table S5.1). All other contrasts (i.e., PET vs SPECT, MRI vs SPECT) were non-significant.

**Table S2.1.** Significant differences identified by contrast analyses

| Contrast | | Region | x | y | x | Volume (mm^3^) |
| --- | --- | --- | --- | --- | --- | --- |
| ***Parkinsonism PET vs MRI*** | 1 | L. Mid. Temporal G.,  L. Sup. Temporal G. | -44.8 | -64.5 | 35.4 | 1440 |
| *PET - MRI*  *Park < HC* | 2 | R. Mid. Temporal G. | 52.4 | -65.4 | 23.5 | 680 |
|  | 3 | R. Inf. Frontal G. | 59.5 | 15.8 | 21.7 | 656 |
|  | 4 | L. Caudate | -14.6 | 11 | 4.2 | 440 |

*Note.* Clusters reported were found to be specific to the PET imaging meta-analysis when compared to the MRI imaging meta-analysis (i.e., PET – MRI). Clusters reported were significant at uncorrected *p* < .001. R. = Right; L. = Left; G. = Gyrus; Mid. = Middle; Sup. = Superior; HC = Healthy controls; MRI = Magnetic Resonance Imaging; PET = Positron Emission Tomography.
