## Supplementary file 3 for "Localization of Abnormal Brain Regions in Parkinsonian Disorders: An ALE Meta-Analysis"

**Supplementary file 3. PRISMA Flowcharts**

**Figure S3.1**

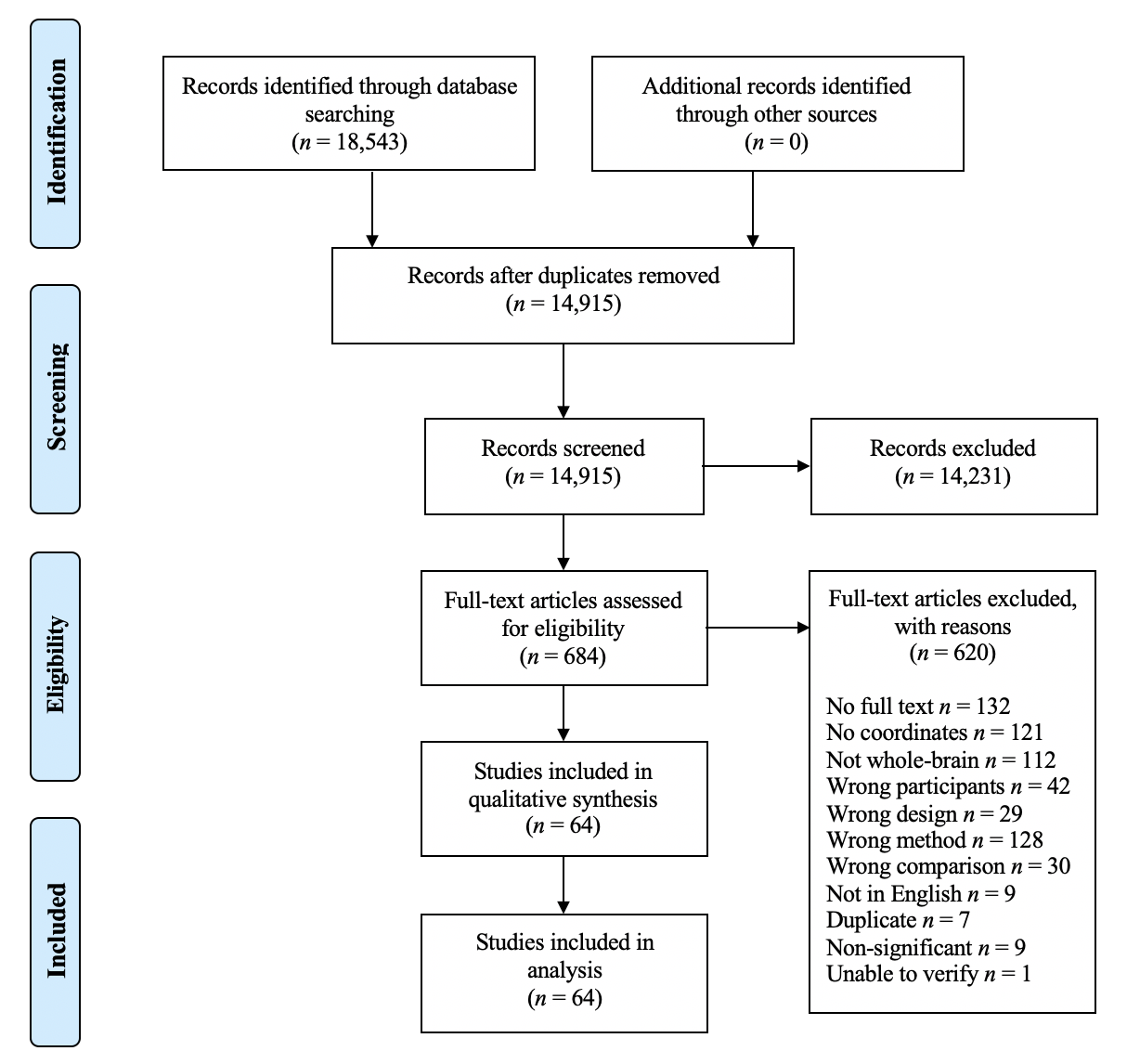
*PRISMA Systematic Literature Search Decision Flowchart for Parkinson’s disease.*

*Note.* The reason ‘Wrong method’ combines: *wrong imaging technique* (113) and *wrong analysis* (15). ‘Duplicate’ combines: *duplicate records* (6) and *duplicated samples* (3) published papers using the same participant cohorts, where possible the largest appropriate sample has been chosen.

**Figure S3.2**

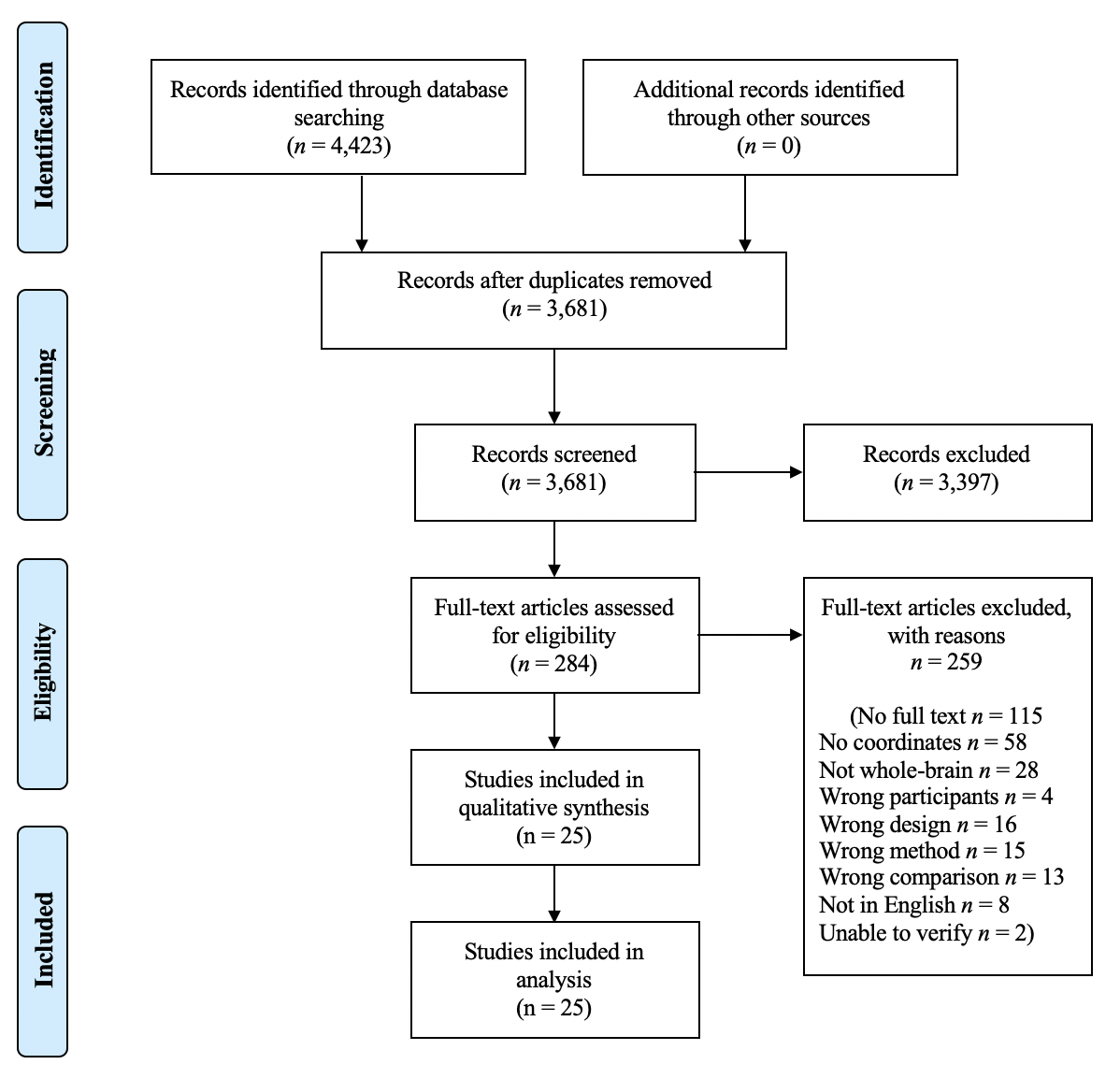
*PRISMA Systematic Literature Search Flowchart for progressive supranuclear palsy*

*Note.* The reason ‘Wrong method’ combines: *Wrong imaging technique* (9); *Results correlated with a task/symptom/effect* (1) and *Wrong analysis* (5).

**Figure S3.3**

*PRISMA Systematic Literature Search Flowchart for corticobasal degeneration*

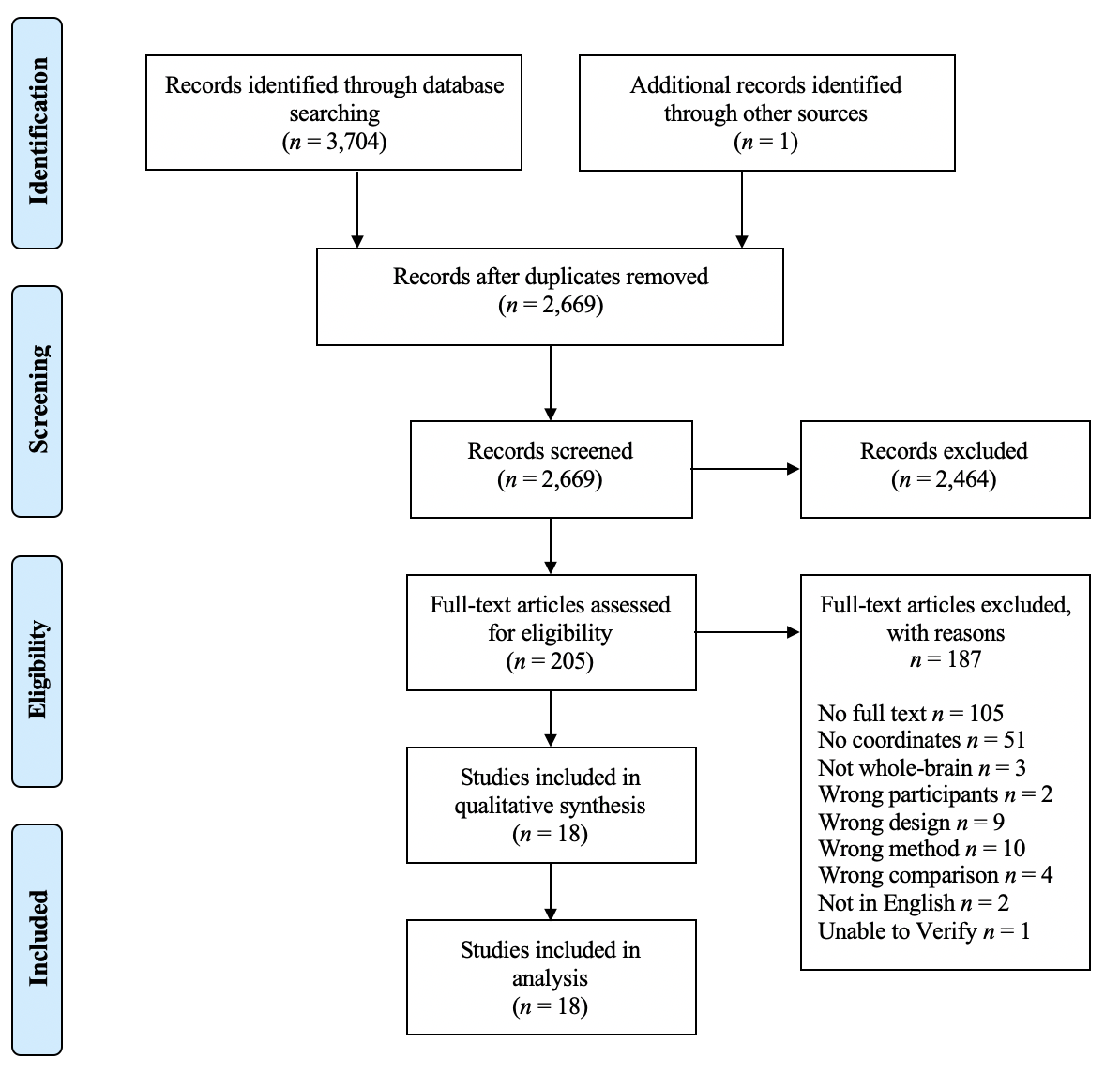

*Note.* The reason ‘Wrong method’ combines: *Wrong imaging technique* (3); *Results correlated with a task/symptom/effect* (7).

**Figure S3.4**

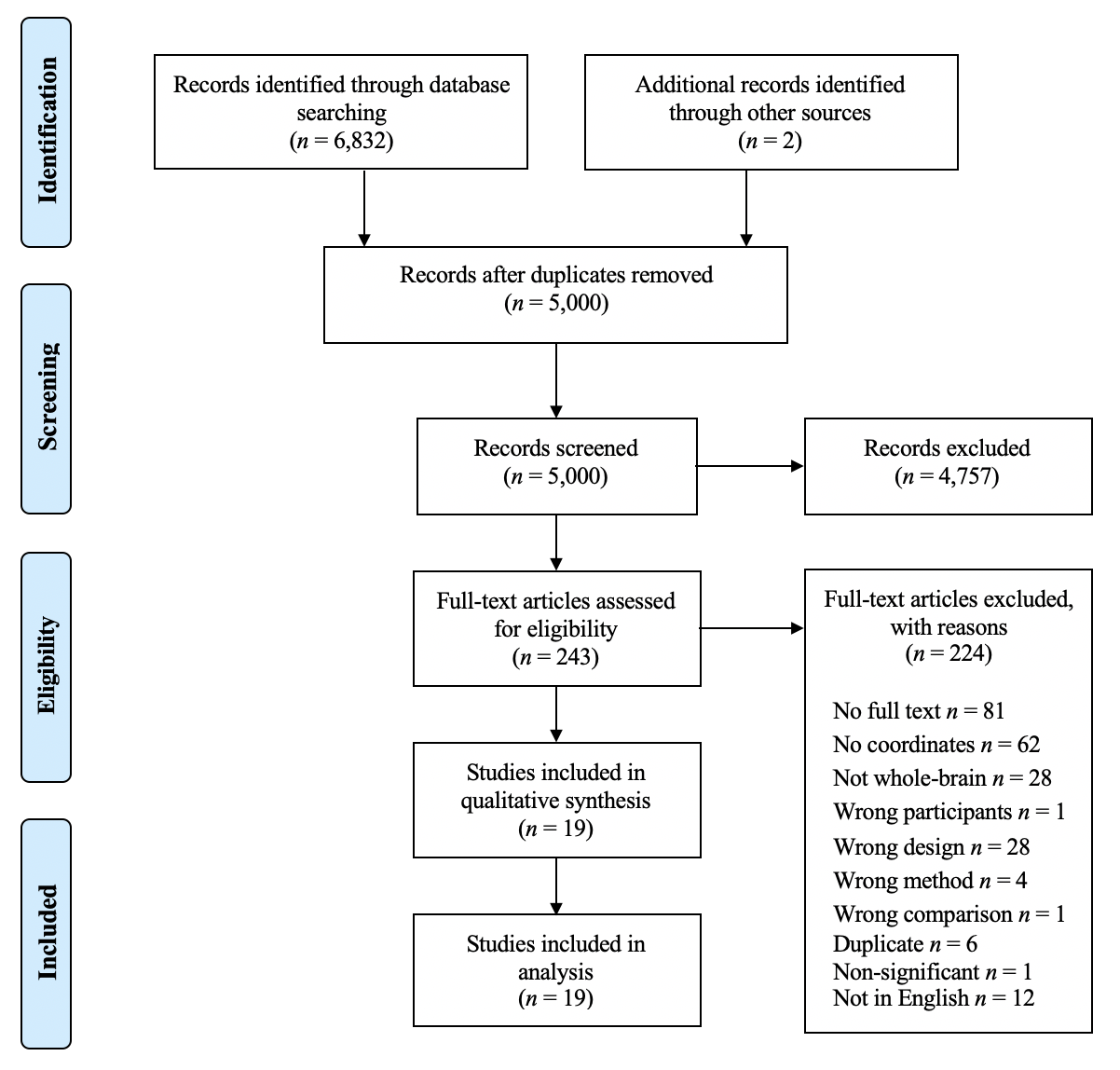
 *PRISMA Systematic Literature Search Flowchart for multiple system atrophy*

*Note.* The reason ‘Duplicate’ combines: *duplicate records* (4) and *duplicate samples* (2) papers that have published using the same participant cohorts, where possible the largest appropriate sample has been chosen.
