## Supplementary file 4 for "Localization of Abnormal Brain Regions in Parkinsonian Disorders: An ALE Meta-Analysis"

**Supplementary file 4. Included Study Characteristics**

**Table S4.1.**

*Included Study Characteristics*

| Study | Diagnosis | Method | *N* Participants | | Mean age (SD) in years | | Diagnostic criteria | Diagnostic Certainty  (if supplied) |
| --- | --- | --- | --- | --- | --- | --- | --- | --- |
|  |  |  | Patients | Controls | Patients | Controls |  |  |
| Abe et al., 2003 | PD | SPECT | 28 | 17 | 67.3 (7.3) | 69.6 (10.2) | UKBB | NS |
| Alzahrani et al., 2016 | PD | MRI – VBM | 40 **PD**  25 **PDA** | 24 | **PD** 66.1 (8.5)  **PDA** 68.7 (8.4) | 62.8 (9.8) | UKBB | NS |
| Burton et al., 2004 | PD | MRI – VBM | 31 **PD**  26 **PDD** | 36 | **PD** 75.2 (5.2)  **PDD** 72.3 (5.2) | 75.1 (6.6) | UKBB | NS |
| Camicioli et al., 2009 | PD | MRI – VBM | 43 | 43 | 70.7 (4.0) | 71.0 (4.5) | UKBB | NS |
| Chen et al., 2017 | PD | MRI - VBM | **PD-NC**: 11  **PD-MCI**: 21 | 21 | **PD-NC:** 59.2 (2.58)  **PD-MCI:** 63.6 (11.16) | 61.1 (8.3) | UKBB | NS |
| Chung et al., 2016 | PD | FDG-PET | **PD-NC**: 13  **PD-MCI**: 11 | 15 | **PD-NC**: 61.5 (7.9)  **PD-MCI**: 68.0 (4.6) | 65.7 (3.7) | UKBB | NS |
| Cilia et al., 2008 | PD | SPECT | **PD**: 40  **PDG:** 11 | 29 | **PD**: 55 (7.0)  **PDG**: 57.4 (5.8) | 56 (6.0) | UKBB | NS |
| Cordato et al., 2005 | PD | MRI – VBM | 17 | 23 | 67.7 (6.7) | 71.5 (7.2) | Gelb Criteria | NS |
| Fioravanti et al., 2015 | PD | MRI - VBM | 20 | 15 | 60.5 (7.7) | 64.6 (4.8) | UKBB | NS |
| Gama et al., 2014 | PD | MRI – VBM | **PD:** 28  **PDVH:** 11 | 10 | **PD**: 65.7 (7.8) **PDVH**: 70.6 (9.1) | 68.1 (7.0) | Unspecified criteria; clinically confirmed with doctor. | NS |
| Gao et al., 2017 | PD | MRI - VBM | **PD:** 23  **PD-MCI:** 23 | 21 | **PD**: 62.4 (7.7)  **PD-MCI**: 65.1 (8.7) | 63.8 (5.4) | UKBB | NS |
| Gerrits et al., 2014 | PD | MRI – VBM | 93 | 46 | 63.0 (10.) | 61.0 (8.0) | UKBB | NS |
| Guimarães et al., 20 | PD | MRI – VBM | 66 | 40 | 57.9 (10.3) | 57.6 (10.8) | UKBB | NS |
| Huang et al., 2013 | PD | FDG-PET | 26 | 12 | 66.5 (1.4) | 67.4 (2.0) | UKBB | NS |
| Li et al., 2017 | PD | MRI-VBM | 366  (from PPMI) | 172 | 62.2 (9.8) | 60.6 (11.4) | Not specified (taken from PPMI) | NS |
| Lin et al., 2013 | PD | MRI – VBM | 10 | 13 | 67.3 (8.8) | 65.3 (11.1) | UPDRS | NS |
| Lyoo et al., 2010 | PD | MRI – CTh | 48 | 56 | 60.2 (9.1) | 60.5 (7.2) | UKBB | NS |
| Potgieser et al., 2014 | PD | MRI – VBM | 77 | 87 | 63.0 (10.5) | 60.1 (7.2) | UKBB | NS |
| Ramírez-Ruizet al., 2007 | PD | MRI – VBM | **PD:** 20  **PDVH:** 18 | 21 | **PD**: NS  **PDVH**: NS | NS | UKBB | NS |
| Summerfield et al., 2005 | PD | MRI – VBM | **PD:** 13  **PDD:** 16 | 13 | **PD**: 72.8 (4.9)  **PDD**: 70.1 (7.9) | 70.1 (7.2) | UKBB | NS |
| Van Laere et al., 2004 | PD | ^99M^TC-ECD SPECT | 81 | 44 | 62.6 (10.2) | 59.2 (11.9) | UKBB | NS |
| Wang et al., 2017 | PD | FDG-PET | **PD:** 15  **PDA:** 13 | 15 | **PD:** 64.1 (9.0)  **PDA**: 68.3 (5.7) | 63.3 (4.6) | UKBB | NS |
| Wilson et al., 2019 | PD | MRI - | **PD-Mild:** 27 **PD-MOD:** 27 **PD-SEV:** 26 | 30 | **PD-Mild**: 57.2 (8.4)  **PD-MOD**: 60.7 (9.1)  **PD-SEV**: 60.1 (10.1) | 60.2 (9.5) | UKBB | NS |
| Yadav et al., 2016 | PD | MRI - | 64 | 46 | 70.3 (6.3) | 71.2 (10.0) | UKBB | NS |
| Zhang et al., 2015 | PD | MRI – VBM | **PD:** 14  **PD-MCI:** 21 | 20 | **PD**: 58.5 (9.2)  **PD-MCI**: 63.8 (8.6) | 59.4 (6.4) | UKBB | NS |
| Hosey et al., 2005 | PD | H_2_^15^O PET | 9 | 9 | 59 (9) | 53 (12) | UKBB | NS |
| Hosokai et al., 2009 | PD | FDG PET | **PDNC**: 27  **PD-MCI:** 13 | 13 | **PD-NC:** 65.7 (5.1)  **PD-MCI:** 67.6 (5.5) | 63.0 (4.6) | UKBB | NS |
| Kostic et al. 2010 | PD | MRI - VBM | **PD total**: 40 | 26 | 66 (NS) range: 50-79 | 63 (NS) range: 48-79 | UKBB | NS |
| Lyoo et al., 2010 | PD | FDG PET | **PD total:** 61  **(PD-NC:** 20  **PD-SA:** 12,  **PD-SN:** 11,  **PD-MD:** 18) | 14 | **PD total:** 64.0 (NS) range 56-68.  **PD-NC:** 62.0 (NS) range: 55.8-73.0.  **PD-SA:** 65.5 (NS) range: 56-71.  **PD-SN**: 57.0 (NS) range: 54-72.  **PD-MD:** 65.5 (NS) range: 60.3-69. | "age matched" | UKBB | NS |
| Pereira et al., 2012 | PD | MRI - VBM | **PD:** 20 | 20 | 64 (9.53) | 59.1 (10.9) | UKBB | NS |
| Teune et al., 2010 | PD | FDG-PET | 20 | 18 | 63 (9) | 56 (14) | MDS Criteria (Litvan et al., 2003) | NS |
| Tir et al., 2009 | PD | MRI - VBM | 19 | 18 | 61.6 (7.6) | 56(14) | MDS Criteria (Litvan et al., 2003); UKBB | NS |
| Juh et al., 2004 | PD | FDG-PET | 8 | 22 | 67.9 (10.7) | 67.8 (14.4) | UKBB | NS |
| Gasca-Salas et al., 2016 | PD | FDG-PET | **PD-MCI:** 12  **PD-MCI+VH:** 9 | 19 | **PD-MCI**: 70.8 (3.4).  **PD-MCI+VH**: 70.7(3.9). | 70.1 (3.1) | UKBB | NS |
| Juh et al., 2005: | PD | FDG-PET | 8 | 22 | 67.9 (10.7). | 67.8 (14.4) | UKBB | Probable |
| Compta et al., 2012 | PD | MRI - VBM | **PD-ND:** 18  **PDD:** 15 | 12 | **PD-ND**: 69 (NS) range: 67.5-76.75 **PDD**:73 (NS) range: 65-78. | 71.5 (NS) range: 67.5-76.75 | UKBB | NS |
| Diez-Cirarda et al., 2015 | PD | MRI - VBM | 37 | 15 | 67.97 (6.17) | 71.5 (67.5-76.75) | UKBB | NS |
| Liang et al., 2016 | PD | MRI - VBM | **PD-ND:** 20  **PD-DEP:** 16 | 21 | **PD-ND**: 56.43 (6.45)  **PD-DEP**: 63.5 (9.87) | 65.07 (7.01) | UKBB | NS |
| Terada et al., 2018 | PD | MRI - VBM | 40 | 10 | 64.7 (8) | 67.6 (3.2) | UKBB | NS |
| Martin et al., 2009 | PD | MRI - VBM | 26 | 14 | 59.8 (7.7) | 56.8 (7.8) | Calne et al., 1992 | NS |
| Naduthota et al., 2017 | PD | MRI - VBM | 72 | 72 | 51.4 (10.6) | 50.8 (10.4) | UKBB | NS |
| Nagano-Saito et al., 2005 | PD | MRI - VBM | **ND-PD*:*** 39  **ND-PD-Adv.:** 19 | 31 | **ND-PD**: 61.8 (8.1)  **ND-PD-Adv.:** 62.6 (7.9) | 63.5 (8.8) | Calne et al., 1992 | NS |
| Nobili et al., 2009 | PD | ^99m^Tc ECD SPECT | **PD-MCI:** 15 | 15 | 71.5 (+/-5.9). | 71.3 (6.1) | Gelb Criteria | NS |
| Pappatå et al., 2011 | PD | FDG-PET | **PD-MCI** 12 | 12 | 64 (5.3) | 62 (6.2) | UKBB | NS |
| Song et al., 2014 | PD | Tc-99m HMPAO SPECT | **TPD**: 33 | 33 | 70.85 (8.65). | 66.94 (5.4) | UKBB | NS |
| Sarro et al., 2013 | PD | MRI - VBM | **PD-MOD:** 14  **PD-SEV:** 12 | 42 | **PD-MOD**: 65 (8);  **PD-SEV:** 65 (7); | 64 (7) | UKBB | NS |
| Pagonabarraga et al., 2014 | PD | MRI - VBM | **PD-NH:** 27  **PD-mH:** 15 | 15 | **PD-NH:** 66.3 (8);  **PD-mH:** 64.1 (9) | 66.8 (8) | UKBB | NS |
| Le Jeune et al., 2010 | PD | FDG-PET | 20 | 13 | 57.9 (9.7) | 53.23 (11.2) | UKBB | NS |
| Lee et al., 2015 | PD | MRI - VBM | **PD-L:** 23  **PD-R:** 23 | 23 | **PD-L**: 60.7 (6.8). **PD-R**: 57.9 (6.5). | 57.9 (6.7) | UKBB | NS |
| Tessitore et al., 2012 | PD | MRI - VBM | **PD-FOG**: 12 | 12 | NS (>45 years) | "age matched" | UKBB | NS |
| Xuan et al., 2019 | PD | MRI - VBM | **EOPD:** 25  **M-LOPD:** 37 | **Young HC**: 23  **Old HC:** 23 | **EOPD**: 49.6 (5.9)  **M**-**LOPD**: 62.4 (5.9) | **Young HC**: 51.3 (6.3)  **Old HC:** 64.2 (6.7) | UKBB | NS |
| Kikuchi et al., 2001 | PD | Tc-99m HMPAO SPECT | **PD-all:** 18 | 11 | 59.1 (11.5) | 62 (9.02) | UKBB | NS |
| Berding et al., 2001 | PD | FDG-PET | 11 | 10 | 56 (10). | 49 (16) | NS | NS |
| Berti et al., 2012; | PD | FDG-PET | 26 | 21 | 65.3 (6.4) | 62.4 (9) | Gelb Criteria | NS |
| Hsu et al., 2007; | PD | Tc-99m HMPAO SPECT | 21 | 11 | 64.4 (8.7). | 60.1 (7.5) | UKBB | NS |
| Imon et al., 1999; | PD | SPECT | 27 | 24 | 65.6 (10). | 61.8 (9) | Ward & Gibb Criteria | NS |
| Danti et al., 2015; | PD | CTh (FreeSurfer) | 28 | 48 | 63.5 (NS) | 58.4 (10) | UKBB | NS |
| Pagonabarraga et al., 2013; | PD | CTh (FreeSurfer) | **PD-NC:** 18  **PD-MCI:** 18 | 18 | **PD-NC:** 60.6 (9). **PD-MCI:** 66.5 (6.7) | 62.3 (7.4) | UKBB | NS |
| Tessitore et al., 2016 | PD | CTh (FreeSurfer) | **PD-NC**: 26 | 18 | 71.5 (4); | 68.2 (4) | UKBB | NS |
| Huang et al., 2016; | PD | CTh (Surface-based morphometry) | **PD-NC** 15  **PD-ICD:** 15 | 24 | **PD-NC:** 63.14 (8); **PD-ICD:** 62.87 (8.6). | 63.54 (6.7) | UKBB | NS |
| Jia et al., 2019; | PD | MRI - VBM | 34 | 45 | **PD-dep:** 59.4 (8.9); **PD-NC:** 59.1 (9.9) | 57 (10) | UKBB | NS |
| Rektorova et al., 2014; | PD | SBM | **PD-NC:** 27  **PD-MCI:**27 | 25 | **PD-NC**: 63.11 (9.27); **PD-MCI**: 62.59 (6.61) | 59.44 (5.77) | UKBB | NS |
| Gasca-Salas et al., 2019: | PD | CTh (FreeSurfer) | **PD-MCI**: 15  **PDD:** 8 | 18 | **PD-MCI:** age: 70.1 (NS), range: 65-69.  **PDD:** 69.6 (NS) range: 66-73.2. | 67.6 (NS) range: 65-69) | UKBB | NS |
| Garcia-Diaz et al., 2017; | PD | CTh (FreeSurfer) | **PD-all:** 36 | 20 | 64.37 (9.97). | 69.15 (7.94) | UKBB | NS |
| Boxer et al., 2006; | PSP | MRI-VBM | 15 | 80 | 70.9 (6.9). | 67.9 (8.6) | NINDS-SPSP | probable |
| Brenneis et al., 2004; | PSP | MRI-VBM | 12 | 12 | 67.5 (6.6) | 60 (5.8) | NINDS-SPSP | probable |
| Cordato et al., 2005 | PSP | MRI-VBM | 21 | 23 | 70.3 (6.4). | 71.5 (7.2) | NINDS-SPSP | 5 definite;  16 NS |
| ^a^Lagarde et al., 2013; | PSP | MRI-VBM | 19^a^ | 18 | 65.9 (6.5). | 67.8 (5.2) | NINDS-SPSP | probable |
| Lehéricy et al., 2010; | PSP | MRI-VBM | 10 | 9 | 66.9 (6.4) | 66.5 (4.8) | NINDS-SPSP | NS |
| Padovani et al., 2006; | PSP | MRI-VBM | 14 | 14 | 73.0 (5.36). | 65.6 (4.1) | NINDS-SPSP | probable |
| Sakurai et al., 2014; | PSP | MRI-VBM | 33 | 32 | 78 (6). | 79 (3). | NINDS-SPSP | 4 possible;  29 probable |
| Takahashi et al., 2011; | PSP | MRI-VBM and FDG-PET | 16 | 20 | 64.6 (6.4). | 64.8 (6.4) | NINDS-SPSP | probable |
| Whitwell et al., 2013; | PSP | MRI-VBM | 16 | 20 | 72.1 (4.6; | 73.9 (6.3) | NINDS-SPSP | probable/definite |
| Agosta et al., 2018; | PSP | CTh (FreeSurfer) | 21 | 36 | 62.9 (6.5). | 64.8 (7.0). | Holinger et al., 2017 | Probable |
| Agosta et al., 2010: | PSP | MRI-VBM | 20 | 24 | 64.9 (NS) range: 53-82 | 63.8 (NS) range: 48-79 | Williams et al., 2005 | 18 probable;  2 possible |
| Ghosh et al., 2012; | PSP | MRI-VBM | 23 | 22 | 71.1 (8.6). | 71.4 (7.6) | MDS Criteria (Litvan et al., 2003) | 9 definite;  14 NS |
| Giordano et al., 2013; | PSP | MRI-VBM | 15 | 15 | 68.91 (1.2). | 65.5 (6.1) | NINDS-SPSP | probable |
| Sandhya et al., 2014; | PSP | MRI-VBM | 10 | 8 | NS | NS | NINDS-SPSP | probable /possible |
| Kimura et al., 2011; | PSP | ^99m^TcECD SPECT | 19 | 17 | 73.2 (8) | 68.8 (10.7) | NINDS-SPSP | probable /definite |
| Park et al., 2009; | PSP | FDG-PET | 14 | 11 | 68.9 (6.4) | 72 (6) | NINDS-SPSP | probable |
| Price et al., 2004; | PSP | MRI-VBM | 12 | 12 | 65.3 (5.8). | 67.4 (4.6) | MDS Criteria (Litvan et al., 2003) | 8 definite;  4 probable |
| Teune et al., 2010; | PSP | FDG-PET | 17 | 18 | 68 (8) | 56 (14) | NINDS-SPSP | 13 probable;  4 possible |
| Varrone et al., 2007; | PSP | ^99m^TcECD-SPCT | 16 | 10 | 67 (6) | 59 (16) | NINDS-SPSP | 12 probable;  4 possible |
| Wang et al., 2015; | PSP | MRI-VBM | 24 | 23 | 64.17 (6.72) | 60.52 (6.47) | NINDS-SPSP | probable /possible |
| Worker et al., 2014; | PSP | CTh (FreeSrufer) | 14 | 19 | 69.4 (7.2) | 63.8 (7.9) | NINDS-SPSP | 13 probable;  1 possible |
| Hosaka et al., 2002; | PSP | MRI-VBM | **12** | 12 | 62.8 (6.0) | 63.8 (7.7) | NINDS-SPSP | probable |
| Ge et al., 2018; | PSP | FDG-PET | **PSP:** 20  **Early stage-PSP**: 14 | **HC**: 20  **early stage HC:** 20 | **PSP**: 65.3 (8.3);  **Early-stage PSP:** 65.6 (8.2) | **HC**: 59.0 (11.4); **Early-stage HC**: 62.5 (6.7) | NINDS-SPSP | probable |
| Juh et al., 2004: | PSP | FDG-PET | 7 | 22 | 67.6 (4.83) | 67.8 (14.4) | Retrospectively clinically diagnosed | NS |
| Lagarde et al., 2015 | PSP | MRI-VBM | 20 | 18 | 65.5 (6.5) | 67.8 (5.2) | NINDS-SPSP | NS |
| Abe et al., (2016). | CBS | ^99m^Tc ECD SPECT | 26 | 26 | 76 (5.3). | 76.6 (5.8) | Mathew et al., 2012 | possible |
| Albrecht et al., (2019). | CBS | MRI-VBM | 25 | 25 | 66.7 (10.1) | 66.2 (10.1) | Otto et al., 2011 | NS |
| Borroni et al., (2008) | CBS | MRI-VBM | 20 | 21 | 62.7 (8) | 65.6 (4.1) | Lang, 1994 | probable |
| Boxer et al., (2006). | CBS | MRI-VBM | 14 | 80 | 64.6 (5.9) | 67.9 (8.6) | NS | Probable |
| Gross et al., (2010). | CBS | MRI-VBM | 20 | 8 | 67.4 (9.8) | 69.4 (3.9) | NS (neurologist assessment) | NS |
| Pardini et al., (2009). | CBS | MRI-VBM | 25 | 14 | 62 (9) | NS | NS (neurologist assessment) | NS |
| Pardini et al., (2019). | CBS | FDG-PET | 14 | 13 | 64.2 (9.5) | 61.5 (6.2) | neurologist assessment and pathological examination (autopsy) | definite CBD (autopsy) |
| Sakurai et al., (2014). | CBS | MRI-VBM | 18 | 32 | 79 (5) | 79 (3) | Lang, 1994 | NS |
| Spotorno et al., (2015). | CBS | MRI - 'PipeDream' analysis | 10 (7 scanned) | 19 | 70 (2) * | NS | Armstrong et al., 2013 | NS |
| Teune et al., (2010). | CBS | FDG-PET | 10 | 18 | 39 (9) | 56 (14) | Mahapatra et al., 2004 | NS |
| ^b^Grossman et al., (2004). | CBS | MRI-VBM | 9 | 25 | 64 (7) | 68.5 (9.4) | ^b^ see note. | NS |
| Halpern et al., (2004). | CBS | MRI-VBM | 13 (5 scanned) | 12 | 66.76 (10.15) * | NS | NS (neurologist assessment) | NS |
| Hosaka et al., (2002). | CBS | FDG-PET | 12 | 12 | 64.8 (6.3) | 63.8 (7.7) | Lang, 1994 | probable |
| Mille et al., (2017). | CBS | FDG-PET | 29 | 20 | 67.4 (7.7) | 68 (7.9) | Armstrong et al., 2013 and  Boeve 2011 | NS |
| Whitwell et al., (2011). | CBS | MRI-VBM | 5 | 20 | 65 (NS) range: 42–69 | 63 (NS) range 50–70 | autopsy | definite |
| Zamboni et al., (2010). | CBS | MRI - VBM | 31 (26 scanned) | 14 | 65.8 (1.5)* | 60.5 (1.9) | Boeve et al., 2003 | NS |
| Misch et al., (2014). | CBS | ^99m^Tc ECD SPECT | 31 | 31 | 68.5 (1.7) | 70 (1.2) | Boeve et al., 2003 | NS |
| Huey et al., 2009 | CBS | MRI-VBM | 48 | 14 | 66 (9) | 60 (6) total:62 | Boeve et al., 2003 | NS |
| Grimaldi et al., (2019). | MSA | FDG-PET | 85 | 60 | median: 66 (range: 60.5–71.5 years) | median: 66 (range: NS) | Gilman et al., 2008 | probable |
| Brenneis et al., (2006). | MSA | MRI-VBM | **MSA-C:** 13 | 13 | 61.3 (6.2) | 60.5 (4.4) | Gilman et al., 1999 | 12 probable; 1 possible |
| Brenneis et al., (2003). | MSA | MRI-VBM | **MSA-P:** 12 | 12 | 62 (6.6) | 60 (5.8) | Gilman et al., 1999 | probable |
| Chang et al., (2009). | MSA | MRI-VBM | **MSA =** 23 [**MSA**-**C** = 10; **MSA**-**P=** 13] | 37 | MSA-C: 57.1 (9.9) MSA-P: 59.8 (8.1) | 55.5 (8.6) | Gilman et al., 2008 | probable |
| Dash et al., (2019). | MSA | MRI-VBM | **MSA:** 26  **[MSA-C** =18; **MSA-P** = 8; 1 undefined] | 25 | 55.7 (5.4) | 55.0 (6.8) | Gilman et al., 2008 | probable |
| El Fakhri et al., (2006). | MSA | ^99m^ Tc-ECD SPECT | 5 | 9 | 66.8 (11.5) | 63.8 (8.1) | Gilman et al., 1999 | probable |
| Fiorenzato et al., (2017). | MSA | MRI-VBM | 72 | 36 | 63.8 (6.8) | 61.6 (7.4) | Gilman et al., 2008 | probable |
| Juh et al., (2005). | MSA | FDG-PET | 11 | 22 | 58.5 (8.4) | 67.8 (14.4) | Quinn Criteria (Wenning et al., 1994) | probable |
| Kimura et al., (2011). | MSA | ^99m^ Tc-ECD SPECT | **MSA-P**: 12 | 17 | 69.3 (7.3) | 68.8 (10.7) | Gilman et al., 1999 | probable |
| Lee et al., (2008). | MSA | FDG-PET | **MSA-C:** 41 | 30 | 56.9 (6.9) | 55.2 (5.8) | Gilman et al., 1999 | probable |
| Minnerop et al., (2010). | MSA | MRI-VBM | 14 | 14 | 61.1 (3.3) | 58.6 (5.1) | Gilman et al., 1999 | NS |
| Shigemoto et al., (2013). | MSA | MRI-VBM | **MSA-P:** 20 | 30 | 62.9 (7.7) | 62.9 (7.7) | Gilman et al., 2008 | 16 probable; 4 possible |
| Specht et al., (2003). | MSA | MRI-VBM | **MSA-C:** 14 | 13 | 59.4 (7.4) | 55.1 (6.9) | Gilman et al., 1999 | 9 probable; 5 possible |
| Teune et al., (2010). | MSA | FDG-PET | 21 | 18 | 64 (10) | 56 (14) | Gilman et al., 2008 | 13 probable MSA-P;  1 probable MSA-C;  7 possible MSA-P |
| Tir et al., (2009). | MSA | MRI-VBM (GM only) | **MSA-P:** 14 | 14 | 63.6 (9.74) | 59.2 (7.6) | Gilman et al., 1999 | probable |
| Tzarouchi et al., (2010). | MSA | MRI-VBM | **MSA-P:** 11 | 11 | 61.9 (11.7) | 64.63 (10.4) | Gilman et al., 2008 | NS |
| Van Laere et al., (2004). | MSA | ^99m^ Tc-ECD SPECT | 15 | 44 | 61.4 (9.2) | 59.2 (11.9) | Quinn Criteria (Wenning et al., 1994) | probable |
| Planetta et al., (2015). | MSA | MRI-VBM | **MSA-P:** 14 | 14 | 64.6 (9) | 61.9 (8.4) | Gilman et al., 2008 | probable |
| Minnerop et al., (2007). | MSA | MRI-VBM | **Total:** 48  **MSA-C**: 32  **MSA-P**: 16 | **Total**: 46  **Subset** (matched to MSA-P): 16 | 61.2 (6) | 58.7 (6.1)  **Subset** 62.3 (4.3) | Gilman et al., 1999 | 8 possible; 24 probable |

*Note*. SD = standard deviation; UKBB = United Kingdom Brain Bank criteria for Parkinson’s disease; PD = Idiopathic Parkinson’s disease; PSP = Progressive Supranuclear Palsy; CBS = Cortiobasal Degeneration Syndrome; MSA = Multiple System Atrophy; HC = Healthy controls; PD-NC = Parkinson’s disease normal cognitive functioning; PD-MCI = Parkinson’s disease mild cognitive impairment; PDD = Parkinson’s disease dementia; PD-mH = Parkinson’s disease -minor hallucinations; PD-NH = Parkinson’s disease-without hallucinations; PD-MILD = Parkinson’s disease mild severity; PD-MOD = Parkinson’s disease moderate severity; PD-SEV = Parkinson’s disease severe severity; PD-FOG = Parkinson’s disease with freezing of gait; EOPD = early-onset Parkinson’s disease; LOPD = late-onset Parkinson’s disease; PD-ICD = Parkinson’s disease impulse control disorder; ND-PD = non-demented Parkinson’s disease; ND-PD-adv = non-demented advanced Parkinson’s disease; PD-ND = Parkinson’s disease without depression; PD-DEP = Parkinson’s disease with depression; PD-MCI+VH = Parkinson’s disease mild cognitive impairment with visual hallucinations; PD-SA = Parkinson’s disease single amnestic; PD-SN = Parkinson’s disease single non-amnestic mild cognitive impairment; PD-MD = Parkinson’s disease multiple domains; PDG = Parkinson’s disease with pathological gambling; PDA = Parkinson’s disease apathy; TPD = Tremor dominant Parkinson’s disease; PD-L = Parkinson’s disease left; PD-R = Parkinson’s disease right; MSA-C = Multiple System Atrophy-Cerebellar; MSA-P = Multiple System Atrophy-Parkinsonism; NS = Not Specified; VBM = voxel-based morphometry; SBM = Source-based morphometry; ^a^ Lagarde PSP - 1 patient did not undergo scanning but age of that patient was included in the M and SD calculations. ^b^Grossman et al. nine patients were given the clinical diagnosis of CBD based on clinical-pathological studies reported in the literature and authors’ own autopsy series (Rinne et al., 1994; Grimes et al., 1999; Riley and Lang 2000; Forman et al., 2002). ^c^Dash et al. was not included in the MSA-P meta-analysis as there were no significant differences reported between MSA-P patients and controls, diagnosis information of the total cohort is provided here. *denotes records where it is not clear whether non-scanned participants were included in the calculation of the demographic averages.

**References for All Included Studies**

References: 1-64 = PD; 65-89 = PSP; 90-107 = CBS; 108-126 = MSA
