## Supplementary file 5 for "Localization of Abnormal Brain Regions in Parkinsonian Disorders: An ALE Meta-Analysis"

**Supplementary file 5. Table of Meta-Analysis Results with Contributing Studies Listed**

**Table S5.1.** *Consistent Regions of Abnormality in Parkinsonian Disorders*

| Cluster | Region | | | x | y | z | Volume (mm^3^) | Convergence % | Contributors |
| --- | --- | --- | --- | --- | --- | --- | --- | --- | --- |
| ***Parkinsonism < HC***  *n* = 125 | | 1 | L. Caudate | -23.8 | 13.1 | 6 | 4096 | 23% | Guimaraes et al 2017; Summerfield et al 2005; Potgiesser et al 2014; Wang et al 2017; Chung et al 2016; Juh et al 2004; Gasca-Salas et al 2016; ^*^Juh et al 2005; Song et al 2014; Xuan et al 2019; Kikuchi et al 2001; Berding et al 2001; Berti et al 2012; Boxer et al 2006; Cordato et al 2005; Agosta et al 2010; Brenneis et al 2004; Wang et al 2015; Ge et al 2018; Whitwell et al 2013; Brenneis et al 2006; Chang et al 2009; El Fakhri et al 2006; Shigemoto et al 2013; Tzarouchi et al 2010; Planetta et al 2015; Minnerop et al., 2007; Pardini et al., 2019 |
|  | | 2 | R. Thalamus | 5.5 | -12.9 | 12.3 | 3240 | 15% | Burton et al 2004; Gasca-Salas et al 2016; Hsu et al 2007; Cordato et al 2005; Lagarde et al 2013; Lehericy et al 2010; Padovani et al 2006; Agosta et al 2010; Takahashi et al 2011; Teune et al 2010; Wang et al 2015; Ge et al 2018; Whitwell et al 2013; Lagarde et al 2015; Tzarouchi et al 2010; Minnerop et al., 2007; Pardini et al., 2019; Pardini et al 2009 |
|  | | 3 | Red nucleus | 2.2 | -20.6 | -7.9 | 1952 | 14% | Boxer et al 2006; Lagarde et al 2013; Lehericy et al 2010; Padovani et al 2006; Agosta et al 2010; Brenneis et al 2004; Takahashi et al 2011; Ghosh et al 2012; Hosaka et al 2002; Teune et al 2010; Wang et al 2015; Kimura et al 2011; Whitwell et al 2013; Juh et al 2004; Minnerop et al., 2010; Shigemoto et al 2013; Abe et al 2016; Borroni et al 2008 |
|  | | 4 | R. Insula | 43.3 | 17.1 | 3.2 | 1168 | 8% | Lin et al 2013; Burton et al 2004; Song et al 2014; Boxer et al 2006; Padovani et al 2006; Agosta et al 2018; Ghosh et al 2012; Wang et al 2015; Ge et al 2018; Albrecht et al 2019 |
|  | | 5 | R. Mid. Frontal G. | 33.3 | 18.2 | 46.7 | 1136 | 10% | ^b^Van Laere et al 2004; Chung et al 2016; Hosokai et al 2009; Gasca-Salas et al 2016; Gasca-Salas et al 2019; Hsu et al 2007; Agosta et al 2018; Worker et al 2014; Grimaldi et al 2019; Minnerop et al., 2010; Grossman et al 2004 |
|  | | 6 | R. Mid. to Inf. Frontal G. | 51.7 | 14.2 | 28.8 | 1048 | 6% | Hosokai et al 2009; ^c^Teune et al 2010; Berti et al 2012; Rektorova et al 2014; ^d^Hosaka et al 2002 |
| ***Parkinsonism < HC*** | | 1 | Brainstem to L. anterior cerebellar lobe | -4.2 | -41.5 | -37.2 | 2144 | 35% | Cilia et al 2008; Hsu et al 2007; Imon et al 1999; Rektorova et al 2014; Teune et al 2010; Ge et al 2018; Specht et al 2003 |
| *n* = 20 | | 2 | L. Thalamus to insula | -27.5 | -17.1 | 11.1 | 1112 | 30% | Hosey et al 2005; Hsu et al 2007; Imon et al 1999; ^e^Teune et al 2010; Ge et al 2018 |
|  | | 3 | R. Mid. Temporal G. | 39.2 | -31.5 | -3 | 864 | 25% | Huang et al 2013; ^f^Teune et al 2010 |
|  | | 4 | R. Hippocampus | 30.5 | -16.9 | -15.6 | 648 | 15% | Cilia et al 2008; Imon et al 1999; Teune et al 2010 |
| ***PD < HC***  *n* = 63 | | 1 | R. Precuneus | 36.3 | -68.8 | 36.3 | 1504 | 17.5% | Fioravanti et al 2015; Chen et al 2017; Gerrits et al 2014; Huang et al 2013; Lyoo et al 2010a; Wilson et al 2019; Kostic et al 2010; Lyoo et al 2010b; Teune et al 2010; Compta et al 2012; Gasca-Salas et al 2019 |
|  | | 2 | L Mid. Temporal G. /Angular G. | -44.5 | -63 | 33.7 | 952 | 11% | Abe et al 2003; Chung et al 2016; Hosey et al 2005; Hosokai et al 2009; Lyoo et al 2010b; Gasca-Salas et al 2016; Gasca-Salas et al 2019 |
|  | | 3 | L Caudate | -16 | 13.6 | 6.9 | 864 | 11% | Summerfield et al 2005; Wang et al 2017; Juh et al 2004; Juh et al 2005; Xuan et al 2019; Berding et al 2001; Berti et al 2012 |
| ***PD > HC***  *n = 12* | | 1 | Brainstem/L. Anterior Cerebellar lobe | -4.3 | -38.7 | -41.1 | 1464 | 33% | Cilia et al 2008; Hsu et al 2007; Rektorova et al 2014; Teune et al 2010 |
|  | | 2 | L Putamen; Globus Pallidus; Thalamus; Claustrum | -23.7 | -14.7 | 2.4 | 1048 | 17% | Hsu et al 2007; Imon et al 1999 |
|  | | 3 | L. Hippocampus (extent to putamen) | 30.3 | -17 | -14.1 | 992 | 25% | Cilia et al 2008; Imon et al 1999; Teune et al 2010 |
| ***APD < HC***  *n* = 62 | | 1 | Bilat. Thalamus/red nucleus | 2.3 | -18.2 | 0.4 | 9960 | 50% | Boxer et al 2006; Cordato et al 2005; Lagarde et al 2013; Lehericy et al 2010; Padovani et al 2006; Sakurai et al 2014; Agosta et al 2010; Brenneis et al 2004; Takahashi et al 2011; Ghosh et al 2012; Hosaka et al 2002; Price et al 2004; Teune et al 2010; Wang et al 2015; Kimura et al 2011; Ge et al 2018; Whitwell et al 2013; Lagarde et al 2015; Juh et al 2004; Brenneis et al 2004; Fioravanti et al 2015; Minnerop et al., 2010; Shigemoto et al 2013; Tzarouchi et al 2010; Minnerop et al., 2007; Abe et al 2016; Borroni et al 2008; Pardini et al., 2009; Grossman et al 2004; Pardini et al., 2019; Huey et al., 2009 |
|  | | 2 | R. Claustrum | 40.2 | 0.5 | -0.6 | 1440 | 12.90 | Agosta et al 2010; Ghosh et al 2012; Sandhya et al 2014; Chang et al., 2009; Kimura et al., 2011; Shigemoto et al., 2013; Minnerop et al., 2007; Teune et al., 2010; |
|  | | 3 | L. Insula/Putamen | -36.7 | 9.3 | 3 | 1384 | 20.97 | Boxer et al 2006; Padovani et al 2006; Agosta et al 2018; Brenneis et al 2004; Wang et al 2015; Kimura et al 2011; Whitwell et al 2013; Chang et al., 2009; Minnerop et al., 2007; Minnerop et al., 2010; Shigemoto et al., 2013; El Fakhri et al., 2006; |
|  | | 4 | L. Caudate | -12 | 7.1 | 12.3 | 1264 | 16.13 | Boxer et al 2006; Cordato et al 2005; Agosta et al 2018; Wang et al 2015; Ge et al 2018; Whitwell et al 2013; Chang et al., 2009; Tzarouchi et al., 2010; Planetta et al 2015; Pardini et al., 2019 |
|  | | 5 | R. Insula | 44.1 | 16.1 | 4.2 | 1240 | 12.90 | Boxer et al 2006; Padovani et al 2006; Agosta et al., 2010; Ghosh et al 2012; Wang et al 2015; Ge et al 2018; Chang et al., 2009; Albrecht et al., 2019 |
|  | | 6 | R. Parahippocampal G. | 17.6 | -12.8 | -14.6 | 1008 | 9.68 | Lagarde et al 2013; Sakurai et al 2014; Agosta et al 2018; Takahashi et al 2011; Chang et al., 2009; Minnerop et al., 2007 |
|  | | 7 | R. Caudate | 14.4 | 10.6 | 10.5 | 824 | 9.68 | ^g^Teune et al 2010; Brenneis et al 2003; Boxer et al 2006; Pardini et al., 2019; Huey et al., 2009 |
|  | | 8 | R. Cingulate G. | 6 | 9.1 | 44.8 | 752 | 9.68 | Lagarde et al 2013; ^g^Teune et al., 2010; Kimura et al 2011; Whitwell et al 2013; Albrecht et al., 2019 |
| **APD > HC** | | 1 | L. Inf. Occipital G. | -40.3 | -81.7 | 1.5 | 688 | 42.85 | ^h^Teune et al., 2010 |
| *n* = 7 | | 2 | R. Mid. Temporal G. | 41.5 | -2.3 | -30.2 | 640 | 42.85 | ^i^Teune et al., 2010; Ge et al., 2018 |
| ***PSP < HC***  *n* = 25 | | 1 | Bilat. Thalamus/Red nucleus | 2 | -15.6 | -0.7 | 11440 | 76% | Boxer et al 2006; Cordato et al 2005; Lagarde et al 2013; Lehericy et al 2010; Padovani et al 2006; Sakurai et al 2014; Agosta et al 2010; Brenneis et al 2004; Takahashi et al 2011; Ghosh et al 2012; Hosaka et al 2002; Price et al 2004; Teune et al 2010; Wang et al 2015; Kimura et al 2011; Ge et al 2018; Whitwell et al 2013; Lagarde et al 2015; Juh et al 2004 |
|  | | 2 | R. Insula | 43.5 | 17.3 | 3.9 | 1392 | 24% | Boxer et al 2006; Padovani et al 2006; Agosta et al 2018; Ghosh et al 2012; Wang et al 2015; Ge et al 2018 |
|  | | 3 | L. Insula/Claustrum | -38 | 11.6 | 2 | 1280 | 20% | Boxer et al 2006; Agosta et al 2018; Brenneis et al 2004; Wang et al 2015; Kimura et al 2011; Whitwell et al 2013; Sandhya et al 2014 |
|  | | 4 | R. Medial Frontal G. and Cingulate G. | 3.5 | 12.3 | 44.4 | 1080 | 20% | Teune et al 2010; Wang et al 2015; Kimura et al 2011; Varrone et al 2007; Whitwell et al 2013 |
|  | | 5 | L. Caudate | -11.4 | 6.4 | 12.6 | 1008 | 16% | Boxer et al 2006; Cordato et al 2005; Agosta et al 2010; Wang et al 2015; Ge et al 2018; Whitwell et al 2013 |
| ***CBS < HC***  *n = 18* | | 1 | R. Caudate | 13.9 | 10.9 | 11.2 | 792 | 28% | Boxer et al., 2006; Pardini et al., 2009; Teune et al., 2010; Pardini et al., 2019; Huey et al., 2009 |
| ***MSA < HC*** | | 1 | R. Putamen/ Claustrum | 36 | -0.5 | 0.3 | 1208 | 37% | Chang et al., 2009; El Fakhri et al., 2006; Fiorenzato et al., 2017; Kimura et al., 2011; Shigemoto et al., 2013; Teune et al., 2010; Minnerop et al., 2007 |
| *n* = 19 | | 2 | Brainstem | 3.6 | -33.1 | -20 | 1080 | 26% | Brenneis et al., 2006; Lee et al., 2008; Minnerop et al., 2010; Shigemoto et al., 2013; Minnerop et al., 2007 |
|  | | 3 | L Putamen | -24.5 | 10.2 | -2.7 | 992 | 21% | Grimaldi et al., 2019; El Fakhri et al., 2006; Shigemoto et al., 2013; Minnerop et al., 2007 |
|  | | 4 | L anterior cerebellar lobe and culmen | -11.3 | -33.5 | -29.6 | 856 | 32% | Brenneis et al., 2006; Lee et al., 2008; Minnerop et al., 2010; Shigemoto et al., 2013; Tzarouchi et al., 2010; Minnerop et al., 2007 |
| ***MSA-C < HC***  *Studies in analysis* = 6 | | 1 | Brainstem | -9.7 | -30.9 | -27.9 | 840 | 4 (66%) | Brenneis et al., 2006; Lee et al., 2008; Specht et al., 2003; Minnerop et al., 2007 |
|  | | 2 | Brainstem | 9.2 | -32.5 | -23.2 | 832 | 4 (66%) | Brenneis et al., 2006; Lee et al., 2008; Specht et al., 2003; Minnerop et al., 2007 |
| ***MSA-P < HC***  *Studies in analysis* = *8* | |  | R. Insula/Claustrum | 39.6 | 0.5 | -1.9 | 1136 | 4 (50%) | Chang et al., 2009; Shigemoto et al., 2013; Minnerop et al., 2007; Kimura et al., 2011 |
|  | | 2 | L. Putamen | -24.3 | 11.9 | -2.6 | 1080 | 4 (50%) | Brenneis et al., 2003; Shigemoto et al., 2013; Tzarouchi et al., 2010; Minnerop et al., 2007 |
|  | | 3 | R. Putamen | 24.6 | 11.5 | -4.7 | 1008 | 4 (50%) | Brenneis et al., 2003; Shigemoto et al., 2013; Tzarouchi et al., 2010; Minnerop et al., 2007 |

*Note* Clusters reported were significant at *p* < .05 family-wise error corrected. Centre of gravity provided in MNI x,y,z coordinates.

^a^Juh et al 2005 contributed coordinates from PD and MSA patient populations. ^b^Van Laere et al 2004 contributed coordinates from PD and MSA patient populations. ^c^Teune et al 2010 contributed coordinates from PD, PSP and CBS patient populations. ^d^Hosaka et al 2002 contributed coordinates from PSP and CBS patient populations. ^e^Teuen et al., 2010 contributed coordinates from both PSP and MSA patient populations. ^f^Teune et al., 2010 contributed coordinates from PD, PSP, MSA and CBS patient populations.^g^Teune et al., 2010 contributed coordinates from PSP and CBS patients. ^h^Teune et al., 2010 contributed coordinates from PSP, MSA and CBS patients. ^i^Teune et al., 2010 contributed coordinates from PSP and CBS patients. R. = Right; L. = Left; G. = Gyrus; Bilat. = Bilateral; PD = Idiopathic Parkinson’s disease; PSP = Progressive Supranuclear Palsy; CBS= Corticobasal Degeneration Syndrome; MSA = Multiple System Atrophy; HC = Healthy controls.
