## Supplementary file 6 for "Localization of Abnormal Brain Regions in Parkinsonian Disorders: An ALE Meta-Analysis"

**Supplementary file 6. Parkinsonian Disorder Meta-Analysis Results at Exploratory Thresholds**

**Table S6.1.** Meta-analysis results at exploratory thresholds

| **Cluster** | | **Region** | **x** | **y** | **z** | **Volume (mm^3^)** | **Convergence %** | **Contributors** |
| --- | --- | --- | --- | --- | --- | --- | --- | --- |
| ***PSP > HC*** | 1 | L. Insula | -35.1 | -17.9 | 25.4 | 360 | 66.6% | Teune et al 2010;  Ge et al 2018 |
| *n* = 3 | 2 | R. Sup. Temporal G. | 39.4 | -0.1 | -31.9 | 264 | 66.6% | Teune et al 2010;  Ge et al 2018 |
| ***MSA > HC*** | 1 | Red Nucleus | 2 | -22 | -16 | 152 | 33.3% | Fiorenzato et al., 2017 |
| *n* = 3 | 2 | R. Globus pallidus/Parahippocampal G. | 20 | -2 | -14 | 152 | 33.3% | Fiorenzato et al., 2017 |
|  | 3 | R. Anterior Cingulate | 6 | 10 | -14 | 152 | 33.3% | Fiorenzato et al., 2017 |
|  | 4 | L. Caudate | -8 | 16 | -14 | 152 | 33.3% | Fiorenzato et al., 2017 |
|  | 5 | L. Anterior cerebellar lobe | -10 | -40 | -4 | 152 | 33.3% | Fiorenzato et al., 2017 |
|  | 6 | L. Thalamus | -6 | -28 | 0 | 152 | 33.3% | Fiorenzato et al., 2017 |
|  | 7 | R. Anterior cerebellar lobe | 6 | -60 | 2 | 152 | 33.3% | Fiorenzato et al., 2017 |
|  | 8 | R. Cuneus | 2 | -70 | 10 | 152 | 33.3% | Fiorenzato et al., 2017 |
|  | 9 | L. Thalamus | -8 | -18 | 10 | 152 | 33.3% | Fiorenzato et al., 2017 |
|  | 10 | R. Thalamus | 6 | -24 | 16 | 152 | 33.3% | Fiorenzato et al., 2017 |

*Note.* Results reported using exploratory thresholds of uncorrected *p* < .001, 100mm^3^ extent minimum. R. = Right; L. = Left; Sup. = Superior; G. = Gyrus; *n* = studies included in the analysis; PSP = Progressive Supranuclear Palsy; MSA = Multiple System Atrophy; HC = healthy controls.
